# Automatic classification of eeg signals, based on image interpretation of spatio-temporal information

**DOI:** 10.1101/2025.02.10.25322019

**Authors:** Or Oxenberg, Michael Fire, Galit Fuhrmann Alpert

## Abstract

Brain-Computer Interface (BCI) applications provide a direct way to map human brain activity onto the control of external devices, without a need for physical movements. These systems, crucial for medical applications and also useful for non-medical applications, predominantly use EEG signals recorded non-invasively, for system control, and require algorithms to translate signals into commands. Traditional BCI applications heavily depend on algorithms tailored to specific behavioral paradigms and on data collection using EEG systems with multiple channels. This complicates usability, comfort, and affordability. Moreover, the limited availability of extensive training datasets limits the development of robust models for classifiying collected data into behavioral intents. To address these challenges, we introduce an end-to-end EEG classification framework that employs a pre-trained Convolutional Neural Network (CNN) and a Transformer, initially designed for image processing, applied here for spatiotemporal representation of EEG data, and combined with a custom developed automated EEG channel selection algorithm to identify the most informative electrodes for the process, thus reducing data dimensionality, and easing subject comfort, along with improved classification performance of EEG data onto subject’s intent. We evaluated our model using two benchmark datasets, the EEGmmidb and the OpenMIIR. We achieved superior performance compared to existing state-of-the-art EEG classification methods, including the commonly used EEGnet. Our results indicate a classification accuracy improvement of 7% on OpenMIIR and 1% on EEGmmidb, reaching averages of 81% and 75%, respectively. Importantly, these improvements were obtained with fewer recording channels and less training data, demonstrating a framework that can support a more efficient approach to BCI tasks in terms of the amount of training data and the simplicity of the required hardware system needed for brain signals. This study not only advances the field of BCI but also suggests a scalable and more affordable framework for BCI applications.

## 1 Introduction

A Brain-Computer Interface (BCI) system is a computer-based framework that records brain signals, analyzes them, and translates them into commands relayed to an output device to support communication between the human brain and external devices [1, 2]. BCI systems are used in various domains, from healthcare to gaming, and are intended for both clinical and healthy individuals [3]. The structure of most BCI systems includes two main characteristic components [4, 5, 6, 7] - a brain signal recording device and an algorithm that transforms the recorded brain signals into commands for controlling devices. An additional optional component provides feedback to users.^1^

The first component of BCI systems is the brain signal recording device. One of the classical and most common recording devices used for BCI systems is the Electroencephalogram (EEG). With an EEG recording device, electrical signals are collected by non-invasive electrodes placed at multiple locations over the scalp and reflect the propagation of underlying neural activity [8]. EEG signals have been shown to be related to various cognitive processes - including movement control, attention, emotion recognition, music perception, and more [9]. Due to the ability to non-invasively detect electric signals related to cognitive processes, EEG has been widely used in research related to neural engineering, neuroscience, and biomedical engineering [10].

The second component of the BCI system is an algorithm that transforms brain signals into commands that can be used to control external devices. This transformation from brain signals to commands often requires feature extraction and classification. The classification process can be either Rule-based, i.e., using predefined rules [11], or by learning patterns in the data using Machine Learning (ML) models, such as support vector machines (SVM) and Random Forest [12, 13].

The feature extraction process in EEG analysis is classically knowledge-based and customized for a particular task by manual definitions that rely heavily on the specific characteristics of the task and based on the domain knowledge of the researcher [14, 15]. For example, for a set of tasks, EEG signals can be classified based on their amplitude around 300 ms post-stimulus, known as P300 events [16, 17, 18]. Knowledge-based feature extraction can result in the manual choice of sub-optimal features, which can diminish the quality of the classification results [13]. For instance, EEG data is often analyzed in the spectral domain and is commonly split into characteristic ranges of Delta (0-4 Hz), Theta (4-7 Hz), Alpha (8–15 Hz), Beta (16–31 Hz), and Gamma (32-70 Hz), which are traditionally linked to documented cognitive functions [19]. Data outside these designated frequency ranges or from non-targeted electrodes is often discarded before analysis. Yet, this might lead to the exclusion of valuable information [20, 21].

Recent advances in deep learning methods offer new ways to process raw or minimally preprocessed input data and extract informative features for the task at hand automatically [22, 23, 24, 25, 26], bypassing the need for the limited approach of manual feature engineering. For example, Schirrmeister et al. [27] demonstrated that deep Convolutional Neural Networks (CNNs), which are neural network architectures specifically designed to process data with a grid-like topology such as images and time-series, can automatically extract spectral power modulations in the alpha, beta, and gamma frequency bands from raw EEG data that is relevant for task decoding. First, the CNN approach achieved an accuracy of 84%, matching the results of manual feature extraction, which achieved an accuracy of 82.1% on the same dataset without the need for expert-driven manual feature engineering. Second, this automatic method highlighted new informative features that were not captured by traditional manual processes, potentially opening up new avenues for improved accuracy as well as new scientific research and advancements. In another research, Ahmed et al. [28] developed a CNN-based model to classify three emotional states using EEG signals from musical stimuli. Their CNN was successful in automatically extracting asymmetry-based features (AsMap), which capture the difference in differential entropy (DE) between all possible pairs of EEG channels. The significance of these AsMap features lies in their ability to represent complex inter-channel relationships that are challenging to capture through traditional manual feature engineering methods. Their automatic features extraction model demonstrated a high classification accuracy of 97%, suggesting somewhat improved performance compared to methods such as differential entropy (DE), which reached an accuracy of 95%.

However, deep learning models are no magic. They require large datasets to avoid data training limitations of overfitting and generalization errors [29, 30]. Specifically, the nature of EEG data collection from human subjects limits the size of collected datasets and imposes data challenges in the BCI field [31, 32]. Subjects cannot be expected to repeat the same mental task repeatedly for extended periods, limiting the amount of recorded data obtainable from the same individual [33]. BCI systems based on a motor imagery paradigm, for example, typically require a training period to adapt the system to each user’s brain, and the long training period may make it impractical due to subjects’ fatigue [34, 35]. In addition, due to large variations in signal characteristics of different subjects, aggregating data from multiple subjects for BCI purposes is also a challenge [36]. Furthermore, in clinical contexts, privacy and proprietary concerns often restrict acquiring large clinical datasets [29, 37]. Consequently, large, publicly available EEG datasets are uncommon [29].

To overcome the challenges of limited EEG data for BCI purposes, researchers have turned to transfer learning techniques for various BCI applications [38, 39, 40]. Transfer learning leverages pre-trained models from other domains, including their architectures and weights, as a foundation for newly trained tasks. This allows the utilization of pre-existing knowledge acquired from extensive training on large datasets, eliminating the need to train new models from scratch [41]. For instance, Yang et al. [40] showcased the benefits of cross-domain transfer learning by applying models trained on EEG tasks to EEG and ECG tasks for neurological applications. Their method achieved high accuracy in seizure prediction—up to 100% for some patients, while significantly reducing training times, a crucial factor for patient care. Additionally, when transferring knowledge from EEG-based models to ECG tasks for sleep staging, they observed a 2.5% improvement in accuracy and over a 50% reduction in training time compared to methods that did not use transfer learning. Despite these encouraging results in terms of accuracy and efficiency, substantial challenges remain in implementing these techniques in real-world clinical settings. One key issue is the use of large numbers of electrodes, 64 and 34 respectively, as reported in the studies above for research settings, that is often not feasible in clinical settings, nor for commercial home use, due to the high cost of multiple channel recording systems. Additionally, practical considerations such as patient comfort, the time required, and the complexity of placing numerous electrodes on the scalp can limit the applicability of these methods [42, 43].

In this study, we developed a non-specific novel classification model that could be theoretically used for any EEG-based BCI tasks. Our framework includes four key components (see Figure 1). First, we transform the data into the spatio-temporal domain to represent it as spectrogram images (one image per recording channel, i.e., electrode). Second, the spectrogram images are used as input for pre-trained classification networks, originally trained on large image datasets unrelated to EEG data, by leveraging transfer learning. We also extract high-level feature vectors from these models to capture essential patterns in the EEG spectrograms. Third, a new channel selection method we developed uses an ensemble model to infer the importance of different electrodes. Subsequently, the framework automatically selects the most significant channels for the classification task, suggesting fewer electrode systems for future recordings. Finally, a Bidirectional Long Short-Term Memory (BiLSTM) is used to extract spatial information, i.e., the relations between the different electrode locations, from the selected feature vectors to obtain the final classification result.

**Figure 1:**
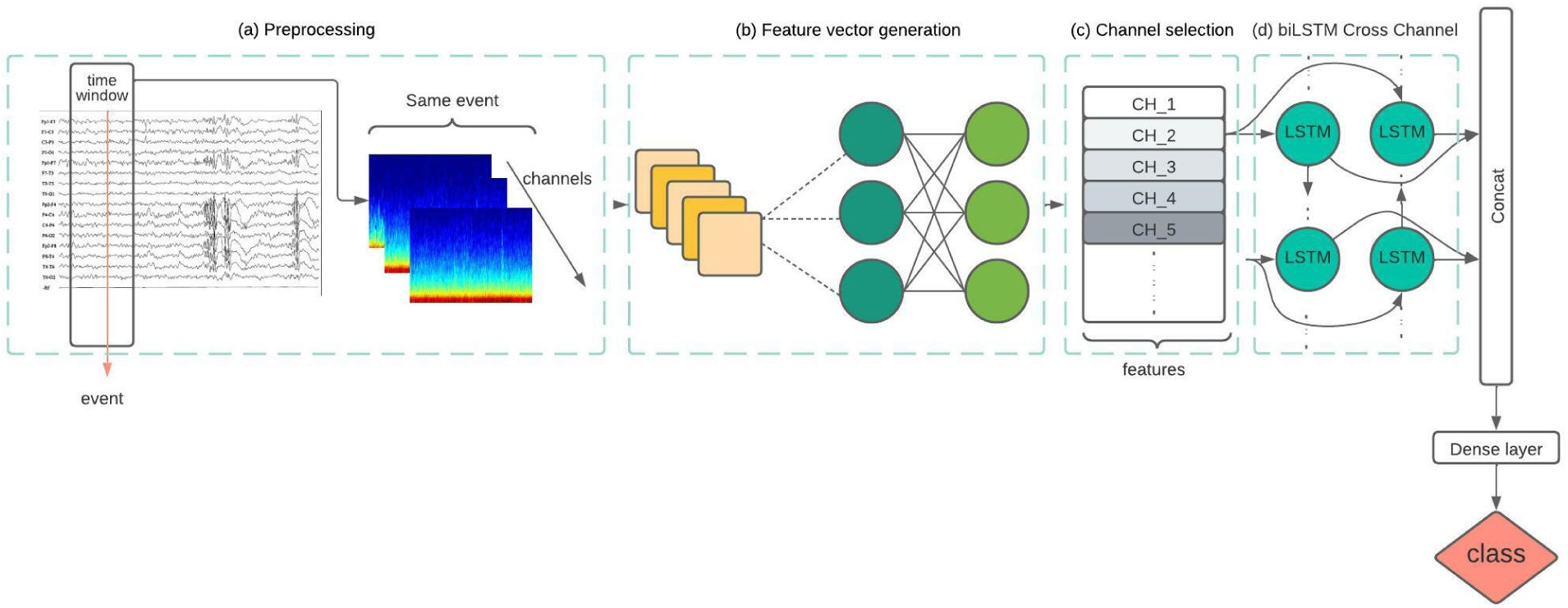
The framework of the EEG-based BCI system consists of four modules: (a) preprocessing the time series EEG data from each channel into spectrogram images; (b) pre-trained CNN to generate feature vectors for each spectrogram image; (c) channel selection method; and (d) classifying the task based on the selected channel feature vector.

To validate the effectiveness and generalizability of our proposed framework, we applied it to two distinct EEG datasets: the Open Music Imagery Information Retrieval (OpenMIIR) dataset and the EEG Motor Movement/Imagery Dataset (EEGmmidb). These datasets represent different EEG paradigms, music imagery, and motor imagery, allowing us to demonstrate that our framework can be applied across a variety of tasks without the need for manual, task-specific adjustments. Additionally, both datasets offer a relatively large number of trials per subject, which, while still smaller than typical datasets used in deep learning, is sufficient to train and evaluate our model effectively.

We compared our model to the foundational classification algorithm for EEG single trial data EEGnet [44], which we consider the benchmark model for this study. We show that our model performs better than the benchmark model for the tested datasets. We further present that it can be trained on relatively small datasets, thus addressing human subject data limitations. Moreover, it automatically selects the most informative electrodes for the classification task. Importantly, this allows the number of electrodes used for classification to be significantly reduced from the multiple-electrode setting (commonly 32,64 or 128 caps for research purposes), as can be seen in both datasets, to as few as three channels only, making it highly applicative for BCI purposes. Another benefit of this procedure is that it reduces noise from irrelevant channels.

## 2 Methods

### 2.1 Classification Framework

We developed an open classification framework that is not task-specific and can be applied to any recorded dataset without requiring manual knowledge-based model design. It could be used for various EEG-based protocols and we demonstrate its performance on two different datasets, as described below.

The framework consists of four steps (see Figure 1):

(a) The raw EEG signals are preprocessed using wavelet transformation to create spectrogram images.
(b) The EEG spectrogram images are being fed into a network architecture that is designed to analyze the entire input simultaneously to efffectively capture spatial features at multiple scales in image data.
(c) The most informative channels are selected an automatic feature selection method.
(d) Inputs from the selected channels are being fed into a BiLSTM architecture that processes data in both forward and backward directions to capture temporal dependencies in sequential data—for final task classification.

We introduce each of these steps in the following subsections.

#### a) Preprocessing

We apply preprocessing for noise removal over the raw EEG signal from each recording electrode. The preprocessing approach is reported by Arunabha Roy [45] and Li et al. [46]. It includes the removal of the DC offset (the linear trend), using a Hamming windowed sinc filter with a low-pass of 20*Hz* and high-pass of 1*Hz*, and finally, removing defective electrodes if provided in the dataset documentation.

Next, we use the MNE Python library, which is a comprehensive open-source software package for processing and analyzing EEG and MEG data^2^ to break the continuous EEG data into discrete epochs according to experimental events. Epoching is a procedure in which specific time windows are extracted from the continuous EEG signal to represent the brain response in a given trial. Time zero is set with respect to the experimental event of interest, as defined specifically for each experimental task at hand. These time windows are called “epochs,” and are usually time-locked with respect to a task-related event. For example, consider an experiment where we are studying the brain’s response to visual stimuli like pictures. The EEG data is continuously recorded as the participant is presented with different images. In this experiment, the event of interest is when a new image is presented to the participant, and we set this as time zero. For example, an epoch can be defined as the EEG data from −200 milliseconds before the onset of the image (the pre-stimulus period) to 800 milliseconds after the onset of the image (the post-stimulus period).

Finally, for each channel and epoch, we employ two complementary signal processing techniques. First, we extract Event-Related Potentials (ERPs), which are voltage fluctuations time-locked to sensory, cognitive, or motor events, by averaging time-locked segments of the EEG signals, allowing us to analyze the neural responses associated with specific events or stimuli. Second, we apply wavelets transformation [47] to map the raw signals in the time domain into spectrogram representations of the signal, a spectral representation in both time and frequency domains. Mammone et al. [48], Ozdemir et al. [49], and Liu et al. [50] presented a similar process in their work.

#### b) Feature Vector Generation

The main idea of the feature vector generation module is to leverage pretrained models to identify high-level patterns in spectrogram images, enabling a more accurate classification of EEG data. By utilizing these models, we aim to extract complex feature representations that capture the intricate characteristics of EEG spectrograms. This approach is particularly beneficial given the limitations in computing power, the time required to train models from scratch, and the scarcity of labeled EEG data. We used two different algorithms to extract feature vectors from the spectrogram images. The first one is usually used in image processing and is based on CNN [48, 49, 50]. The other is based on the attention mechanism, usually used in natural language processing, and shown to be relevant to computer vision [51]. In both cases, we used pretrained models due to their ability to leverage complex feature representations without the need for extensive training. This approach allows more efficient and accurate extraction of relevant features from our spectrogram images, particularly given the limitations in both computing power and time required to train models from scratch.

The specific models used in our study are the Inception-v3 model, which is a large pre-trained CNN for classification, and the DINOv2 (DIstillation with NO labels) model, which is based on transformers, a type of neural network architecture designed for handling sequential data and capturing long-range dependencies [52]. Each spectrogram image was represented by a feature vector of size 2048 when using the Inception-v3 model, and a feature vector of size 1024 when using the DINOv2 model. The intuition for using the Inception-v3 model relies on the research by Raghu et al. [53], who evaluated ten different pre-trained CNN architectures to identify the most effective network for seizure-type classification using EEG data. These architectures varied in depth, layer composition, and overall design, representing a range of state-of-the-art CNN models. Raghu et al. found that the Inception-v3 model achieved the highest classification accuracy among the tested architectures. The DINOv2 model was chosen because it excels in generating high-quality embeddings from images without the need for labeled data [51]. This characteristic is particularly advantageous in our context, where labeled EEG data can be scarce or expensive to obtain.

#### c) Channel Selection

To improve the final classifier’s accuracy and provide a faster and more cost-effective framework, we developed an approach to reduce the number of features fed into the classifier.

We use the Wrapper method [54] as our feature selection process. This approach uses a specific classification model to assess the accuracy of different candidate feature subsets, selecting the one that performs best. Feature selection with the wrapper approach has been used in the literature to reduce the dimensionality of EEG data in EEG classification pipelines, achieving higher classification accuracy and reduced computational complexity [55, 56, 57]. To be more specific, we use the Wrapper method with minor changes. Considering all possible combinations of features across all channels would result in a high-dimensional feature space (number of channels multiplied by 2048 features per vector), making the feature selection process computationally infeasible. To address this, we assume that the data from each channel can be initially evaluated independently for the purpose of feature selection. This means we treat each channel separately and assess its individual contribution to the classification performance based on its feature vector.

For the classification model, we use eXtreme Gradient Boosting (XGBoost) [58], a powerful ensemble learning algorithm based on decision trees designed for speed and performance. It has been shown that specifically for EEG, XGBoost achieves the highest classification accuracy compared to other machine learning classification models such as Bayesian, KNN, Decision Tree, and Random Forest as evaluated on diverse datasets from various domains within EEG classification [59, 60, 61]. We apply the XGBoost model to each channel separately, using feature vectors generated by either the Inception-v3 model or DINOv2. Finally, we calculated the average accuracy for the XGBoost per channel by applying 10-fold cross-validation. After sorting the average accuracy, we selected the top *N* channels for the next phase, the BiLSTM network. *N* is chosen in a grid search process described below. The Wrapper method mentioned above, combined with the XGBoost classifier, automatically selects the *N* most informative channels for the the final classification.

#### d) BiLSTM: Final Task Classification

The final classification step uses the dimensionality reduced matrix as input, representing data from single epochs (see Section 2.1). This matrix has dimensions determined by the number of top *N* channels, as selected in the previous step (Figure 1, bidirectional LSTM) and the size of the feature vector extracted for each channel. This matrix is fed as input to the BiLSTM model. The BiLSTM model is an extension of the Long Short-Term Memory (LSTM) network [62], a type of neural network specialized in recognizing patterns in sequential data such as text, genomes, handwriting, or spoken words [63, 64]. Unlike traditional neural networks that assume inputs are independent, LSTMs utilize internal memory states to process sequences of inputs, making them ideal for tasks where context or order is significant. The bidirectional nature of BiL- STM allows information to flow in both forward and backward directions, enabling the model to capture dependencies from both past and future contexts in the sequence [65]. By structuring the input data as matrix of channels and features, we treat the channels within each epoch as a sequence. This allows the BiLSTM to learn inter-channel relationships by processing the sequence of channel feature vectors, ultimately predicting the classification label for each epoch.^3^ Our adoption of BiLSTM aligns with commonly used conventional hybrid deep learning frameworks, which combine CNN and LSTM networks [66, 67, 68, 69]. In these hybrid architectures, CNNs are used to extract spatial features, while LSTMs capture temporal dependencies. Our framework is built on these type of hybrid architectures. From this point onward, we will refer to this approach as Transfer-BiLSTM. By integrating these two types of networks, we aim to effectively model both the spatial and temporal characteristics of EEG signals for improved classification performance.

### 2.2 Hyper Parameter Tuning

We perform parameter optimization through a simple grid search over the parameters space to enhance the performance of the entire pipeline, a method widely used in brain-computer interface machine learning algorithms [70, 71]. We based the optimization on the performance of the validation set, a set that our framework did not train on. It is important to note that we train a model per subject separately, and different hyperparameters per subject are selected using the training process. The specific parameters being tuned include the learning rate, batch size, epochs, dropout rate, and dropout, among others. A summary of the tuned parameters is described in the Table S6. This hyper-parameter tuning process was applied not only to our primary model but also to the alternative models used for benchmarking, ensuring a fair comparison across different algorithmic approaches.

### 2.3 Datasets

One of the objectives of this study was to evaluate the performance of our classification framework on different EEG paradigms to demonstrate that the framework can be used for a wide range of EEG experimental paradigms without the need for manual task-specific settings. Hence, we test our framework on two EEG paradigms from different fields of interest. We chose datasets containing a relatively large number of trials per subject, in order to allow feasible analysis and facilitate robust evaluation of our deep learning based framework. These are relatively large datasets for EEG data, containing 240 and 44 relevant trials per subject in OpenMIIR and EEGmmidb, respectively. Nevertheless, it should be noted that they are relatively small datasets compared to datasets typically used for deep learning models.

#### a) Open Music Imagery Information Retrieval (OpenMIIR)

The Open Music Imagery Information Retrieval (OpenMIIR) is a well-known public dataset published by the University of Western Ontario [72]. It includes EEG data from 14 participants recorded by BioSemi Active-Two system recording with 64 EEG channels at 512 Hz. It consists of 12 recorded stimuli in randomized order with four conditions, a total of 240 trials (12 stimuli x 4 conditions x 5 blocks) per subject. The four conditions are as follows:

1. The subject listens to a piece of music after a cue is played.
2. The subject imagines a piece of music after a cue is played.
3. The subject imagines a piece of music without a cue.
4. The subject imagines a piece of music without a cue and then presses one of two buttons, indicating whether they believe they have correctly imagined the stimulus.

**Figure 2:**
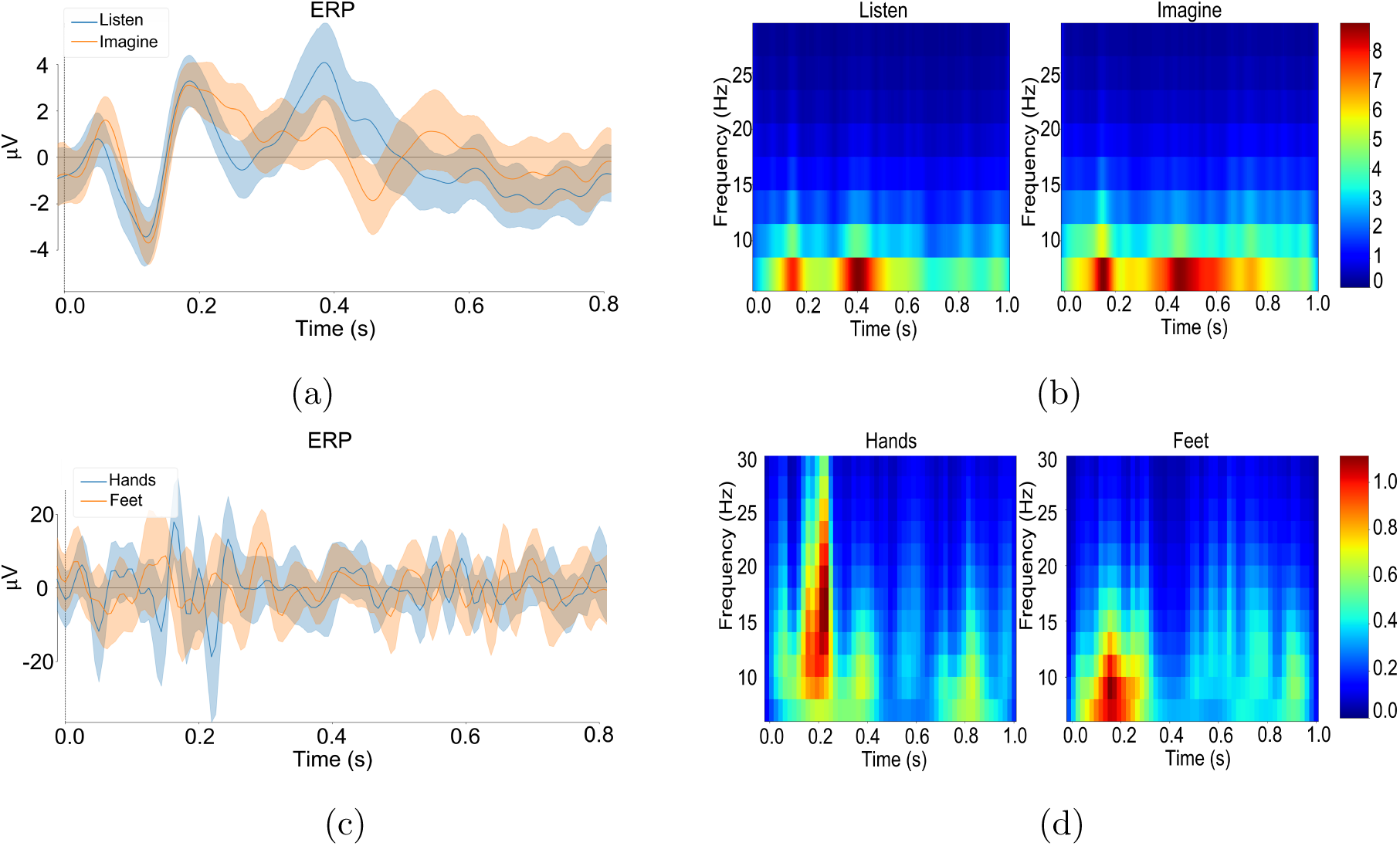
ERP and spectrogram for one subject and one channel for each dataset. (a) + (b) OpenMIIR - both images calculated over channel C3 and subject 1. (c) + (d) EEGmmidb - both images calculated over channel AF3 and subject 1.

In this work, we restrict our analysis to the first two conditions. Using the cued stimuli allowed us to directly compare the perception and imagination of music.

#### b) EEG Motor Movement/Imagery Dataset (EEGmmidb)

The EEG Motor Movement/Imagery Dataset (EEGmmidb) is a collection of EEG recordings focusing on motor movement and imagery tasks. This dataset is designed to capture brain activity patterns associated with performing different motor tasks, such as imagining specific hand movements or performing actual hand movements. The data was recorded using the BCI2000 64-channel system.^4^ Each subject performed 14 experimental runs: baseline runs (one with eyes open and the other with eyes closed) and runs of each of the four following tasks:

1. A cue appears on the screen’s left or right side and the subject opens and closes the corresponding fist until the cue disappears. The subject relaxes between trials.
2. Same as the previous task, only now the subject is asked to only imagine opening and closing his fist.
3. A cue appears and the subject opens and closes both fists until the cue disappears, depending on the location of the visual cue
4. Like the previous task, the subject is now asked to imagine opening and closing fists and feet.

We chose to focus on motor imagery tasks (tasks 2 and 4), as they have proven to be highly valuable in the field of BCIs due to their potential in operating external devices solely through mental activity [73, 74, 75]. In particular, we chose the fourth task focusing on motor imagery. Task four is an expansion of task two, as it encompasses both fists and feet, whereas task two focuses only on fists. Therefore, we expect similar results using the second motor imagery task.

### 2.4 Benchmark models

We tested our framework with comparison to four alternative benchmark models: SVM (Support Vector Machine), EEGNet, XGBoost, and Decision Tree. SVM is a supervised learning algorithm that finds the optimal hyperplane to classify data points into distinct categories [76]. SVM is commonly used in EEG-based BCIs, particularly in the context of binary classifiers [77, 78, 79]. EEGnet [44], a pipeline for robust EEG classification, is a compact CNN for EEG-based BCIs that demonstrates generalization across various BCI paradigms despite limited data availability and produces interpretable features. EEGNet is a robust comparative baseline due to its relatively simple yet effective algorithm. Additionally, it is one of the algorithms with accessible code, making it convenient to use as a benchmark over our datasets.

### 2.5 Evaluation

We applied 10-fold cross-validation to evaluate each model’s performance using multiple metrics to provide a comprehensive assessment, including accuracy which is the percent of correct trial classification and convergence rates which we define as the number of training epochs needed to converge. With a lower convergence rate, we are able to train the model in a shorter period of time. To assess the performance of our framework, we tested two variations of the framework, one utilizing Inception-v3 and the other employing DINOv2 for the feature vector generation model. We conducted a comparative analysis of both accuracy and convergence rates compared to the four benchmark models presented in Section 2.4. Furthermore, we modified the number of channels used for each model and measured the accuracy to determine the impact.

## 3 Results

In this section we present the performance of the proposed framework over the two datasets, in terms of EEG signal classification accuracy. Other metrics such as AUC, F1, and recall, were also applied, and are detailed in Table S1. We also test the effect of using subsets of channels on the classification accuracy and convergence rates.

First, we used the wavelets transformation method to map raw EEG signals to spectrogram images. For OpenMIIR, we created a total of 3658 spectrogram images per subject: 59 for each epoch related to the event IMAGINE or LISTEN, for each subject and each channel. We use eight subjects and 62 channels, so we generated an overall of 29,264 images. For the EEGmmidb datatset, we created spectrogram images for 22 epochs, one second after the cue event, involving left fist movements and 22 events involving right fist movements, for each subject and each channel. The database contains information on ten subjects and 64 channels, so we generated 2816 spectrogram images per subject, and a total of 28,160 images.

Figure 2 presents the averaged spectrogram and event-related potential (ERP) over epochs of a representative subject in a specific channel for each dataset. The channel displayed in the figure is the best-performing channel based on our channel selection model. We present two visualizations of the different events, one showing their amplitude (raw EEG signals) and the second showing their frequency (spectrogram images). This figure illustrates how the temporal information can help to differentiate events using spectrogram images compared to ERP.

We used the generated spectrogram images as input for the DINOv2 and Inception-v3 network models. We extracted feature vectors of sizes 1024 and 2048, respectively. For the EEGmmidb dataset, we generated 28,160 feature vectors, and for the OpenMIIR data, we generated 29,264 vectors. This allows us to evaluate the framework’s performance on datasets of varying sizes, demonstrating its applicability to both small and large datasets.

Next, we consider the channel selection process. To this end, we first measured the classification accuracy for each channel classified separately using the XGBoost model for each event (see Appendix Tables S2, S3, S4 and S5). Then, we selected the top *N* channels sorted by the accuracy for each subject, where *N* is one of the following tested numbers of channels - 3, 5, 9, 15, 30, and 64 channels. Our results in Figures 3 and 4, demonstrate how the number of channels used for model training affects its performance, as measured by accuracy on the test set. We can observe that across different participants (P1-P10), accuracy generally increases with the number of channels up to a certain point, after which it stabilizes or fluctuates slightly. Each subplot shows three algorithms - Transfer-BiLSTM while one leveraging DINOv2 and antoher Inception-v3, and EEGNet, with accuracy plotted against the number of channels selected for each participant. The shaded areas represent the variance across multiple validation folds. Transfer-BiLSTM with DINOv2 tends to perform better across most participants, especially when fewer channels are used. As shown, the channel selection process allows us to drastically reduce the number of channels while preserving high classification performance.

**Figure 3:**
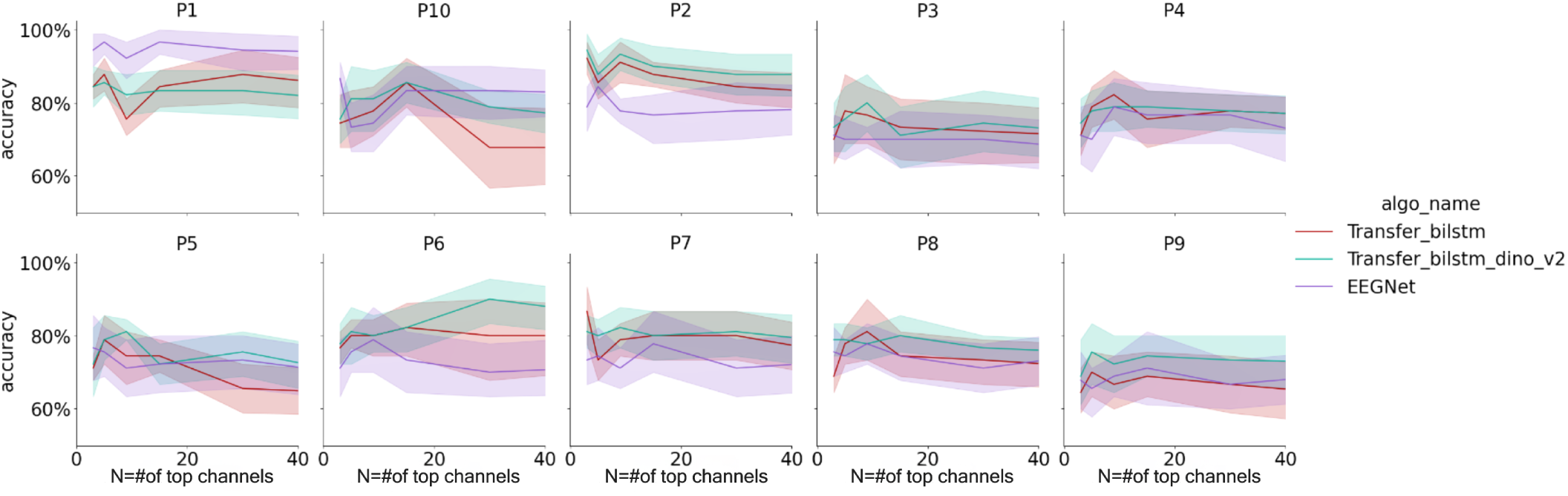
The average accuracy over 10 fold for each subjects, presented in sub-figures, and each model, on the top 3, 5, 9, 15, 30, all channels, calculated over EEGmmidb.

**Figure 4:**
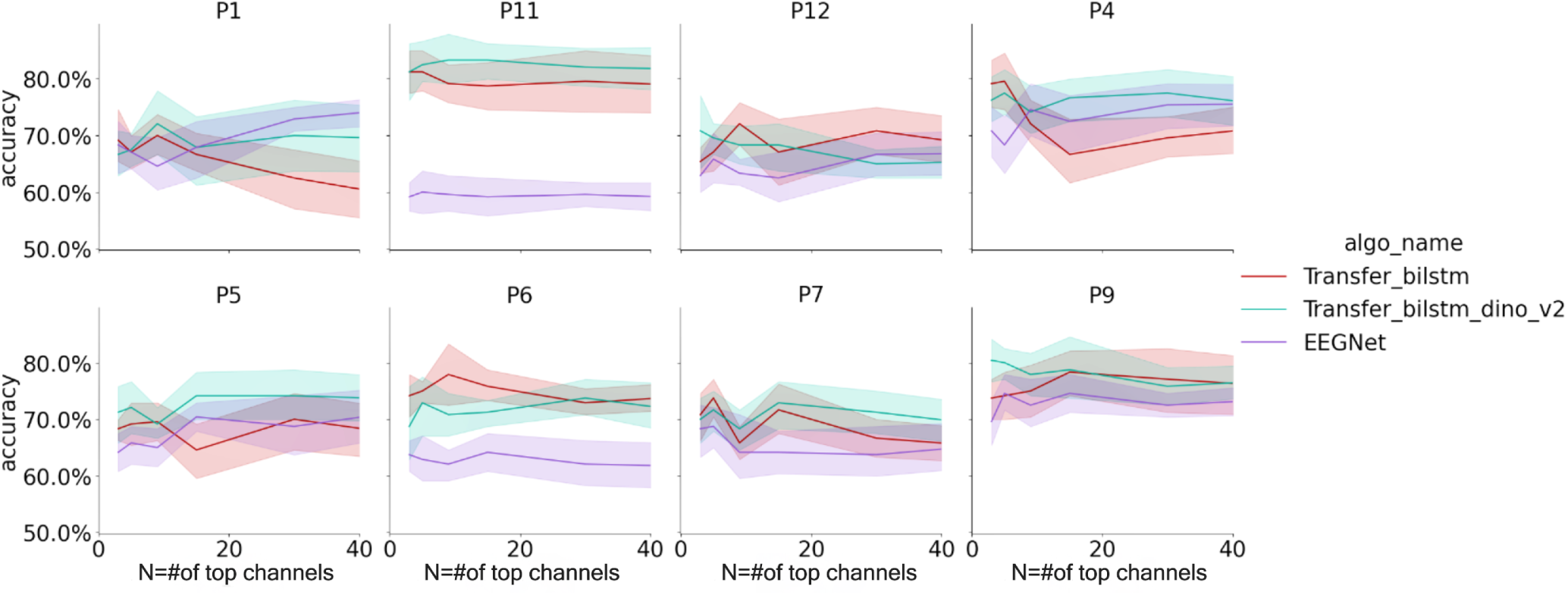
The accuracy over 10 fold for each subjects, presented in sub-figures, and each model, on the top 3, 5, 9, 15, 30, 64 channels, calculated over OpenMIIR.

In the final step of our classification framework, we used the feature vectors generated previously as input which we propagate on to a BiLSTM model and compared its performance with the four benchmark models: EEGnet, SVM classifier, XGBoost, and Decision tree. For the evaluation metric we calculated the average accuracy over the different 10-fold cross validation for each subject. In addition, we evaluate the performance using AUC, F1 score, and recall, as detailed in S1.

Figures 5 and 6 illustrate the results for EEGmmidb and OpenMIIR datasets, respectively. For both datasets, we test our model considering different subset of channels:

1. Using only the top 3 channels for each subject.
2. Using the top *N* channels selected by the channel selection process. Each subject on each dataset has a different *N* selected channels as described in appendix table S6.
3. Using all available channels - 64 channels for EEGmmidb and 62 channels for OpenMIIR.

**Figure 5:**
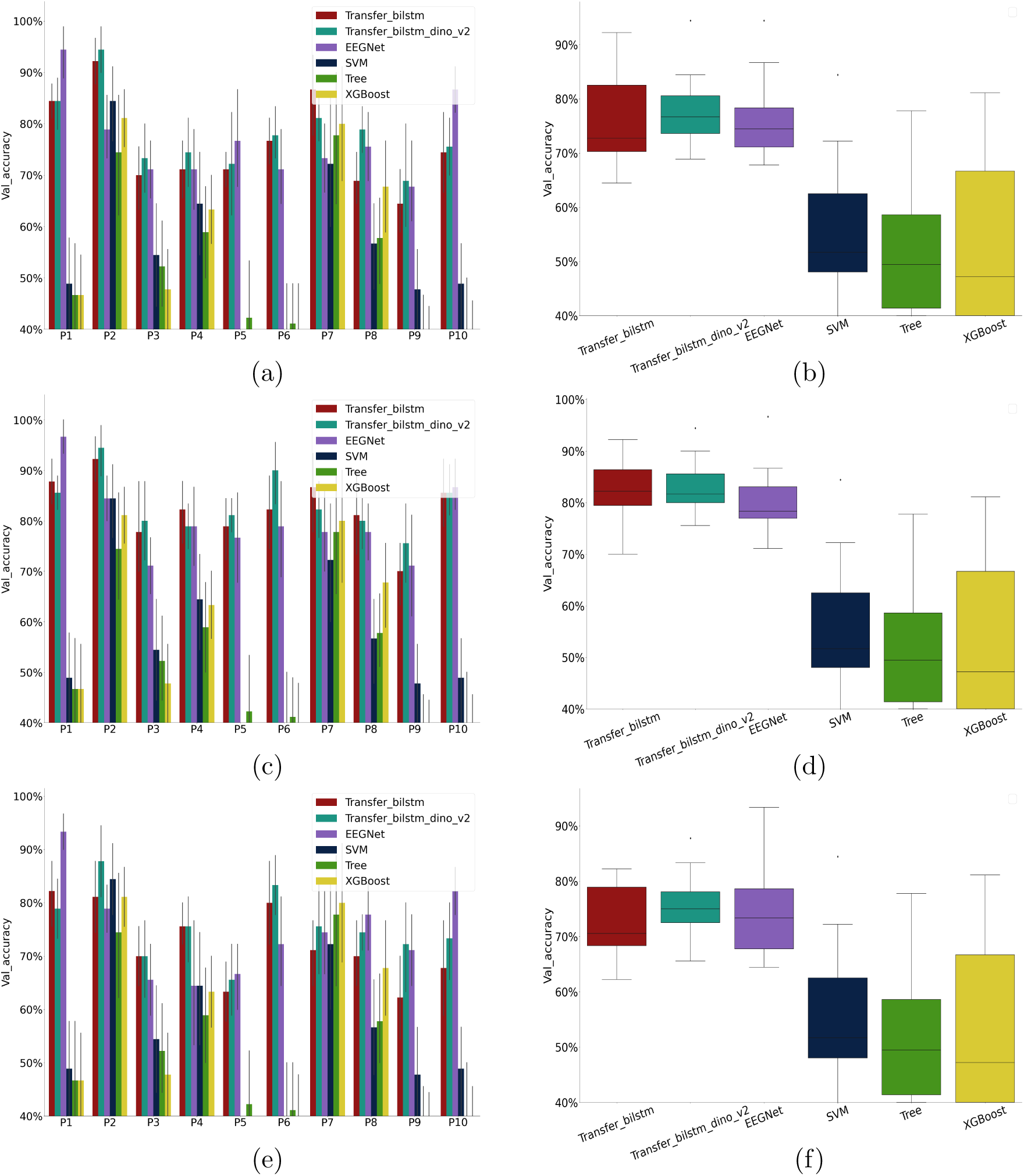
Accuracy comparisons over the test set for **EEGmmidb** dataset. For each number of channels used in the classification we create two types of graphs: mean accuracy over folds for each subject and mean accuracy over all subjects: (a) + (b) three top channels. (c)+(d) best result in the grid search for each model (e)+(f) all channels.

**Figure 6:**
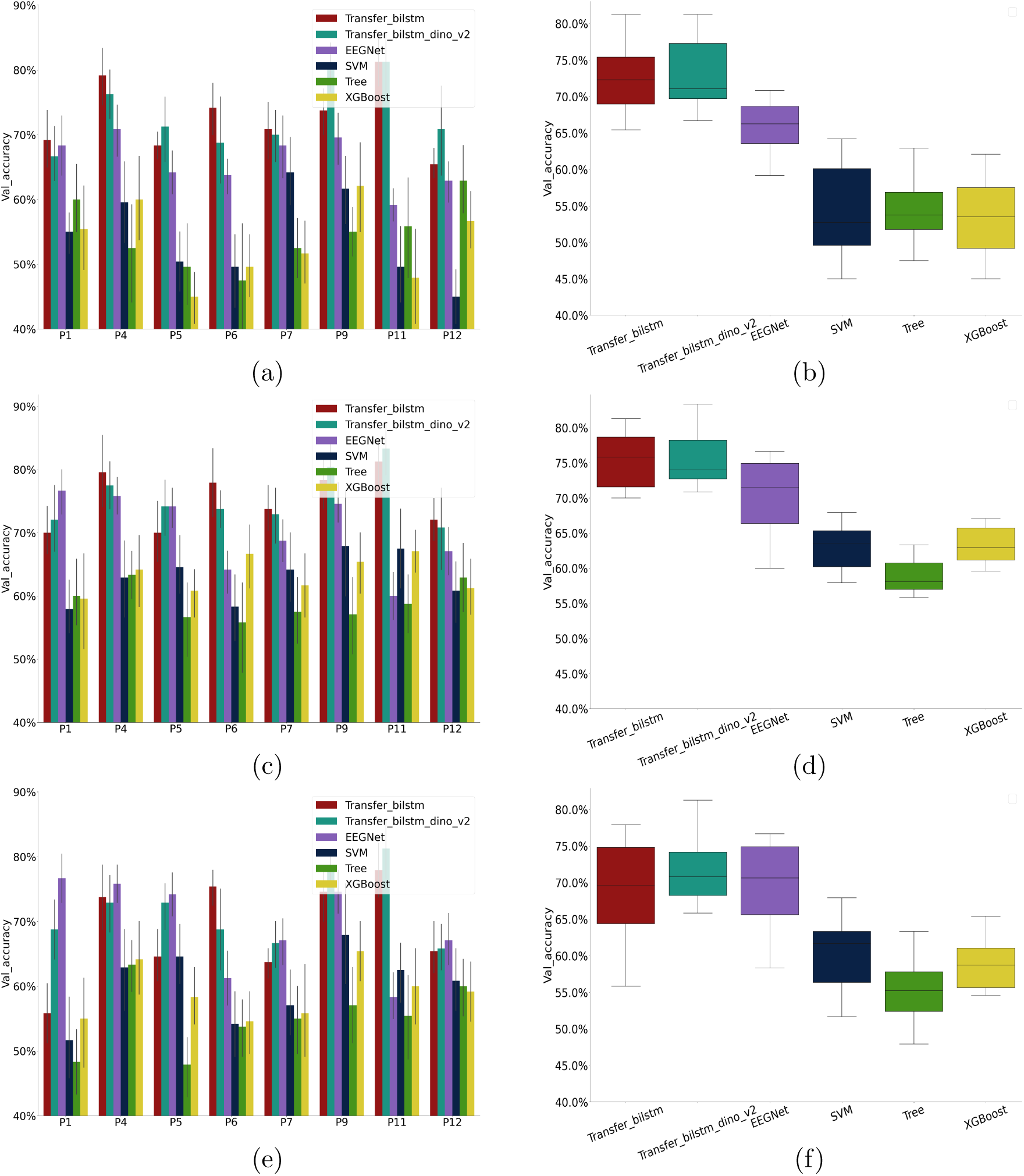
Accuracy comparisons over the test-set for **OpenMIIR** dataset. for each number of channels used in the classification we create two types of graphs: mean accuracy for each subject over folds and mean accuracy over all subjects: (a) + (b) three top channels. (c)+(d) best result in the grid search for each model (e)+(f) all channels.

For each dataset, the left-side graphs in the Figures 5 and 6 (a,c,e) depict the mean accuracy over folds for each subject and the right-side graphs (b,d,f) summarize the mean accuracy averaged over all subjects. The results for each of the models is shown in different colors.

Figures 5a, and 5b, show the average accuracy over the top three channels. Transfer-BiLSTM models, particularly those utilizing DINOv2, consistently outperform the benchmarks, achieving higher mean accuracy across almost all subjects. The only exception was in subject #1, #5 and #10 where EEGnet model achieved higher accuracy compared to our Transfer-BiLSTM models. Figures 5c, and 5d present the best results for each model using the top *N* channels selected for each subject, specifically for the Transfer-BiLSTM models. For example, for subject #10, the best result for the Transfer-BiLSTM model was achieved by using the top 15 channels, while the best result for the EEGnet was achieved by using the top three channels. These again highlight the strong performance of the Transfer-BiLSTM models. Figures 5e and 5f show the results using all channels with Transfer-BiLSTM models showing the highest accuracy, except for subjects #10, #1 and #7.

For the OpenMIIR dataset we observe slightly different results. Figures 6a, and 6b show that Transfer-BiLSTM models perform consistently better for all subjects, with higher mean accuracy over all validation folds. However, the models that do not leverage DinoV2 exhibited better performance. Figures 6c and 6d showed similar results as Figures 6a, and 6b with the only exception of subject #1 where EEGnet model achieved higher accuracy compared to our Transfer-BiLSTM models. Figures 6e and 6f present that using all channels with Transfer-BiLSTM models gives the highest accuracy compare to the benchmarks model, with the exception of subject #1 where EEGnet model again achieved higher accuracy.

Finally we calculated the convergence rate, defined as the number of training epochs needed for the classifier to converge to optimal and stable accuracy. Figures 7 and 8 show the classification accuracy on the training data as a function of the number of epochs used for training. When applying Transfer-BiLSTM with both Inception-v3 and DINOv2 on the EEGmmidb dataset, we can see convergence around 10 epochs, with high accuracy of approximately 90% over the train-set. EEGnet yielded similar results over the training set. For OpenMIIR, the highest accuracy along with the lowest convergence rate were achieved by the Transfer-BiLSTM framework levraging Inception-v3, followed by DINOv2, and finally EEGNet, with a similar convergence rate. Overall, Transfer-BiLSTM had the lowest convergence rate over both datasets.

**Figure 7:**
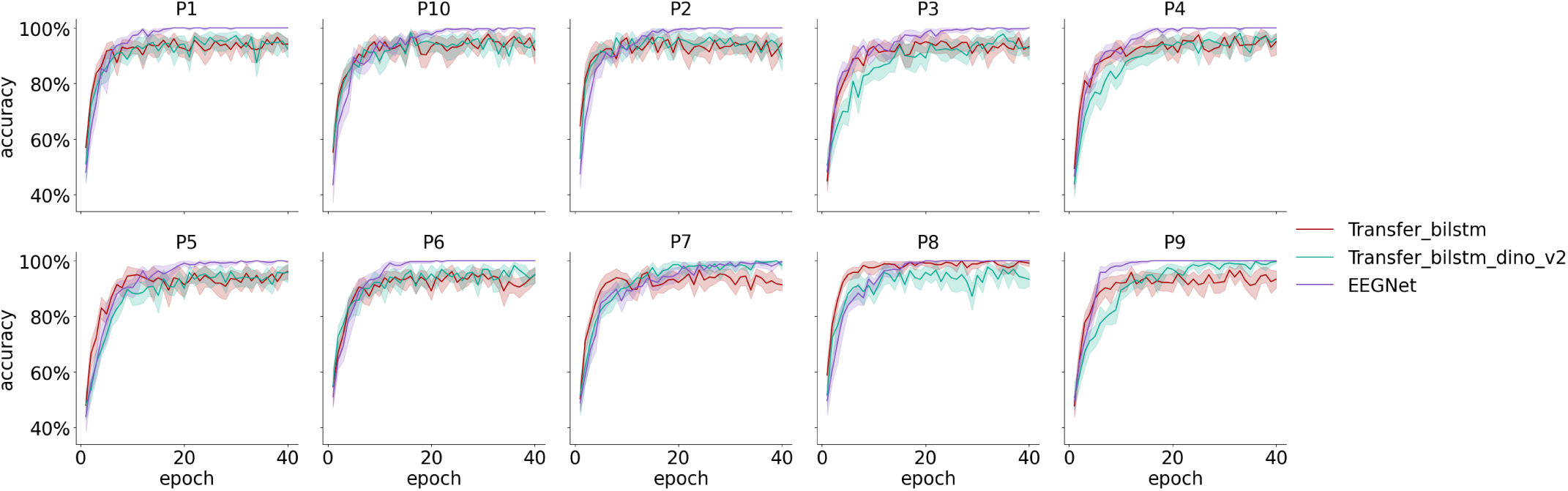
The mean accuracy over folds for each model over each epoch is calculated on a 10-fold basis. A different subject from EEGmmidb is shown in each of the sub-figures.

**Figure 8:**
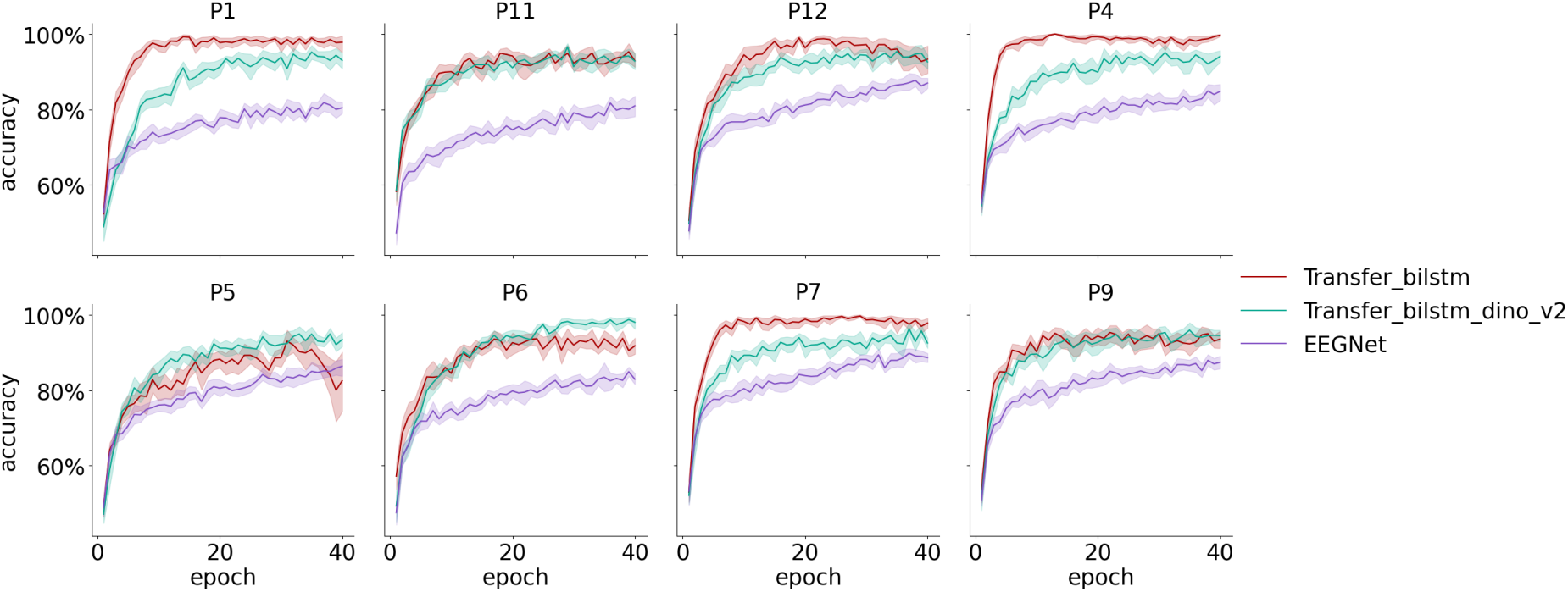
The mean accuracy over folds for each model over each epoch is calculated on a 10-fold basis. A different subject from OpenMIIR is shown in each of the sub-figures.

## 4 Discussion

In this study, we present a robust pipeline for EEG classification that effectively addresses major challenges in EEG data analysis: handling the limitations of small EEG datasets for BCI purposes, reducing the reliance on a large number of electrodes, and enabling automatic feature extraction for improved efficiency and accuracy.

We test it over two dataset, OpenMIIR and EEGmmidb. For OpenMIIR, our Transfer-BiLSTM Inception-v3 model reached an average accuracy of 75%, outperforming EEGnet by 7% (see Figure 6d). For EEGmmidb, our Transfer-BiLSTM using DINOv2 model achieved an average accuracy of 81%, surpassing EEGnet by 1% (see Figure 5d). This improvement is noteworthy given the already high performance of existing models on this dataset. This substantial improvement underscores the versatility of our approach across different EEG paradigms.

Importantly, besides offering improved classification performance, our framework also uses a channel selection method that enables a maintained high accuracy even when drastically reducing the number of channels used.

Our use of spectrogram images as input allowed the models to learn from both frequency and power spaces, enhancing their versatility across various EEG-based BCI paradigms. This approach, combined with transfer learning from pre-trained models like Inception-v3 and DINOv2, effectively addressed the challenge of limited EEG data. By leveraging complex feature representations learned from vast image datasets, we were able to extract meaningful features from our relatively small EEG datasets. We showed that we achieve comparable and even greater performance using as few as three channels, showing that our method can filter out noise from electrodes that are not informative for the task. This capability has profound implications for the practicality of EEG-based applications, potentially enabling the development of more user-friendly and cost-effective EEG systems.

The transfer learning approach also significantly reduced the convergence rate of training by half the trials needed (from around 20 epochs to 10 epochs). This faster convergence is crucial for scaling up to larger numbers of subjects, as it allows for more efficient model training and adaptation. By requiring fewer trials, the approach reduces the duration and intensity of data collection sessions, thereby improving participant comfort and compliance, which are critical factors for real-world applications and larger-scale studies.

However, our framework is not without limitations. It is prone to overfitting, even when regularization is applied. This susceptibility to overfitting likely stems from the large number of parameters that need to be optimized, a common challenge in deep learning models applied to relatively small datasets. To mitigate this, we conducted extensive hyperparameter searches, but future work could explore more advanced regularization techniques or data augmentation strategies to enrich the spectrogram images.

Our framework’s ability to achieve high accuracy with fewer channels and less training data represents a significant advancement in EEG-based BCIs. The generic nature of our approach, in a way that is not task-specific, can easily be applied to any behavioral paradigm, as shown in this study for two very different tasks - the motor imagery and the music imagery. Particularly, in the feature extraction stage, our framework allows for future improvements to be incorporated as newer embedding methods become available. This modularity ensures that our framework can evolve with advancements in deep learning and signal processing techniques.

## 5 Conclusion

We presented robust pipeline for EEG classification, which effectively address major challenges in EEG data analysis, including data scarcity, handling of numerous electrodes, and model agnosticism. By transforming EEG data into spectrogram images and utilizing pre-trained models, as Inception-v3 and DINOv2, we achieved high classification accuracy. Our channel selection method, combined with XGBoost, ensures the framework remains effective even with a reduced number of electrodes, making it more practical for real-world applications.

The proposed framework outperforms existing models such as EEGnet, SVM, Decision trees, and XGBoost, demonstrating its potential for enhancing Brain-Computer Interface (BCI) systems. Our findings indicate that the integration of deep learning models and innovative channel selection methods can significantly improve the practicality and accuracy of EEG-based BCIs, paving the way for more accessible and efficient neurotechnological applications.

The ability to classify using a small number of channels simplifies the use of EEG in practical settings by enabling the development of dedicated small EEG caps with fewer electrodes. These caps are easier to install, more user-friendly, and cheaper to mass-produce, thus enhancing the accessibility and feasibility of EEG-based applications.

In future work will plan to focus on incorporating advanced regularization methods and extending our approach to multi-label classification tasks. In addition, both datasets contain information solely on healthy subjects. We aspire that our framework will be able to generalize on targeted patients (i.e., severely disabled persons), and future work should focus on training and testing our work on people with ALS or other disabilities for brain-wave based communication and control.

## Data Availability

All data produced are available online at openmiir and physionet

https://openmiir.github.io/

https://archive.physionet.org/pn4/eegmmidb/

## A Methods

### A.1 Evaluation Metrics

We provide a comprehensive overview of the evaluation metrics used to assess the performance of our XGBoost and BiLSTM models. The models were subjected to 10-fold cross-validation, ensuring robust and reliable performance estimates. We used metrics like accuracy, f1 score, recall and precision. In addition we use area under the curve which measures the ability of the model to distinguish between classes for multiple thresholds using the model predicted probability. Below are the detailed description and results for each metric.

Table S1 presents a detailed breakdown of our classification model’s performance across various dimensions.

**Table S1:**
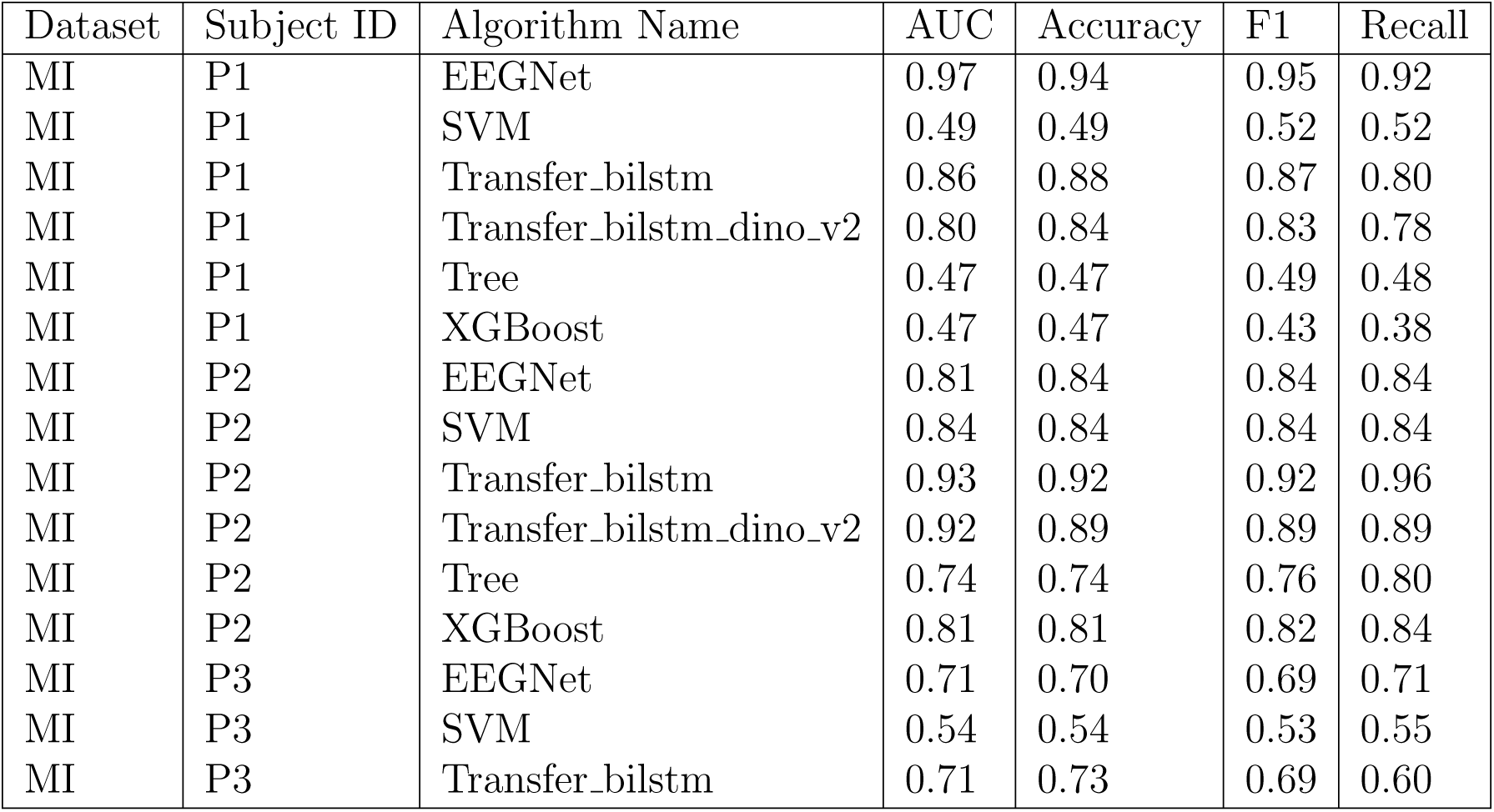
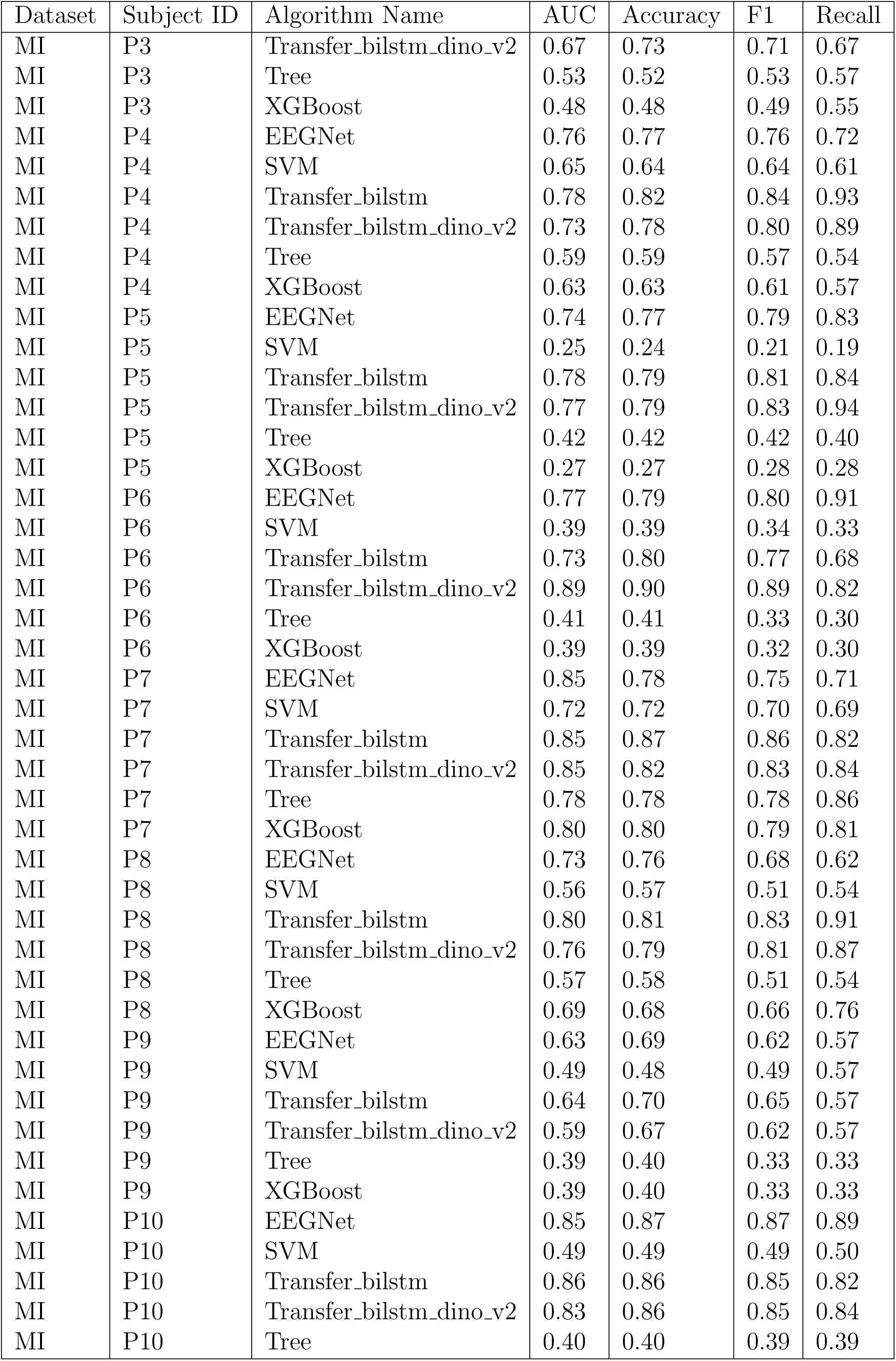
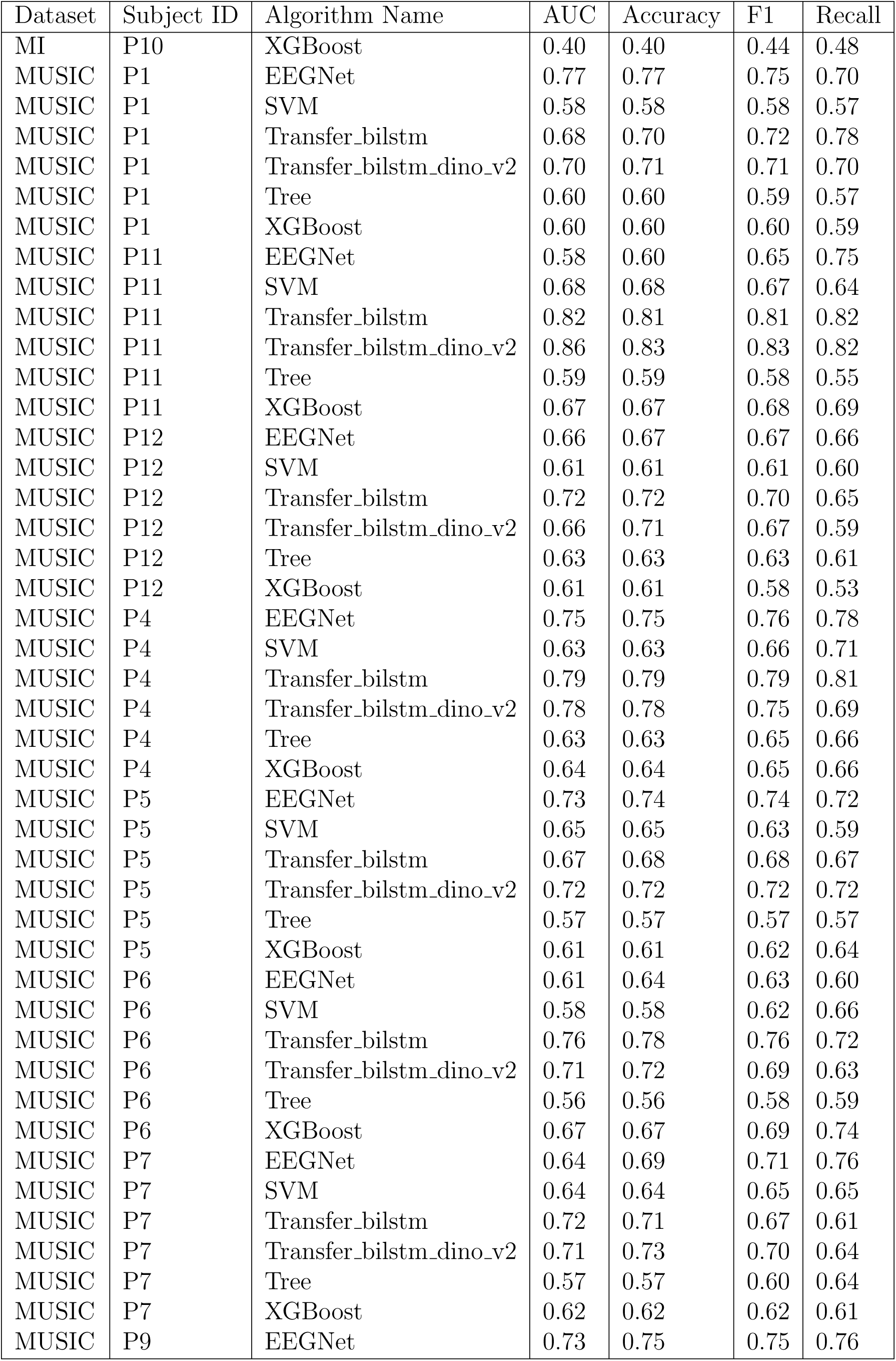
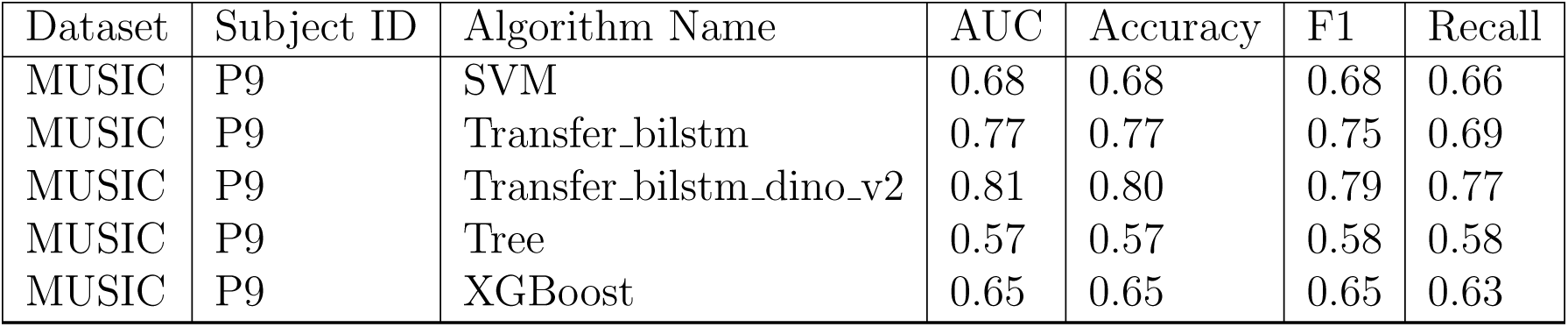
Performance metrics across datasets, subjects, and algorithms.

## B Feature Vector Generation

To leverage the power of deep learning, we transformed our raw EEG signals into spectrogram images, which were then used as input to the DINOv2 and Inception-v3 models. These models are well-known for their feature extraction capabilities in image processing tasks.

- **DINOv2:** For this model, we extracted feature vectors of size 1024 from each spectrogram image.
- **Inception-v3:** This model provided feature vectors of size 2048 from the spectrogram images.

These feature vectors represent the transformed and high-dimensional embeddings of the original EEG signals, providing rich information for further analysis.

- **EEGmmidb Dataset:** We generated a total of 1408 feature vectors using both models.
- **OpenMIIR Dataset:** A significantly larger dataset, we generated 29,264 feature vectors.

## C Channel Selection Process

The next crucial step in our framework is channel selection. This process is vital to identify the most informative channels that contribute significantly to the classification task. We followed a systematic approach to evaluate the impact of different channels. We trained and evaluated the classification pipeline multiple times, each time with a different subset of channels. The primary objective was to measure the classification accuracy for each channel when classified separately using the XGBoost model.

## D Classification Accuracy for Each Event

The classification accuracy for each channel and each event was meticulously measured and documented. This granular analysis allows us to understand the contribution of each channel in the classification task. Table S2, S3, S4 and S5 provides a summary of the classification accuracy for the top three channels for each subject, algorithm, and dataset.

### D.1 Hyper Parameter Tuning

To maximize the efficiency and effectiveness of our entire classification pipeline we performed parameter optimization through a simple grid search over the following parameters:

- **Learning rate:** Regulates the step size at which the optimizer makes updates to the weights during training.
- **Batch size:** Number of samples that will be propagated through the BiLSTM network.
- **Epochs:** One entire transit of the training data through the BiLSTM network.
- **Units:** Number of neurons in a particular layer that impacts the model’s capacity and complexity.
- **Dropout:** A regularization technique for reducing overfitting in neural networks by randomly dropping out a certain percentage of neurons and their connections during training.
- **Lambda:** Controls the “absolute value of magnitude” of the coefficient as a penalty term to the loss function. Specifically, we used the Least Absolute Shrinkage and Selection Operator (Lasso) regression.
- **Number of Top channels (N):** In our pipeline, we sort the channels by their prediction accuracy given by the XGBoost model. We found the optimized N by comparison of various numbers of selected top channels.

We performed hyperparameter tuning for each subject individually to optimize the performance of our transfer bilstm dino v2 and transfer bilstm Inception-v3 models. Table S6 presents the best parameters that were chosen during the hyperparameter tuning process for each subject in each model.

**Table S2:** Summary of the classification accuracy for the top three channels for each subject, and dataset for EEGmmidb using transfer_bilstm.

| Dataset | Algorithm Name | Subject ID | Channel Name | Mean Accuracy |
| --- | --- | --- | --- | --- |
| mi | Transfer_bilstm | p1 | AF3 | 0.69 |
| mi | Transfer_bilstm | p1 | POz | 0.69 |
| mi | Transfer_bilstm | p1 | T10 | 0.67 |
| mi | Transfer_bilstm | p10 | Cz | 0.67 |
| mi | Transfer_bilstm | p10 | FT7 | 0.6 |
| mi | Transfer_bilstm | p10 | P6 | 0.58 |
| mi | Transfer_bilstm | p2 | C5 | 0.77 |
| mi | Transfer_bilstm | p2 | P6 | 0.71 |
| mi | Transfer_bilstm | p2 | CPz | 0.66 |
| mi | Transfer_bilstm | p3 | AF3 | 0.66 |
| mi | Transfer_bilstm | p3 | F7 | 0.63 |
| mi | Transfer_bilstm | p3 | FC1 | 0.61 |
| mi | Transfer_bilstm | p4 | F7 | 0.72 |
| mi | Transfer_bilstm | p4 | FC4 | 0.67 |
| mi | Transfer_bilstm | p4 | CP5 | 0.62 |
| mi | Transfer_bilstm | p5 | Fpz | 0.64 |
| mi | Transfer_bilstm | p5 | PO4 | 0.6 |
| mi | Transfer_bilstm | p5 | F8 | 0.6 |
| mi | Transfer_bilstm | p6 | F3 | 0.68 |
| mi | Transfer_bilstm | p6 | POz | 0.67 |
| mi | Transfer_bilstm | p6 | C3 | 0.64 |
| mi | Transfer_bilstm | p7 | FT8 | 0.68 |
| mi | Transfer_bilstm | p7 | F8 | 0.66 |
| mi | Transfer_bilstm | p7 | AFz | 0.66 |
| mi | Transfer_bilstm | p8 | Fpz | 0.68 |
| mi | Transfer_bilstm | p8 | PO8 | 0.66 |
| mi | Transfer_bilstm | p8 | FC6 | 0.64 |
| mi | Transfer_bilstm | p9 | Fz | 0.67 |
| mi | Transfer_bilstm | p9 | PO8 | 0.64 |
| mi | Transfer_bilstm | p9 | FC4 | 0.64 |

**Table S3:**
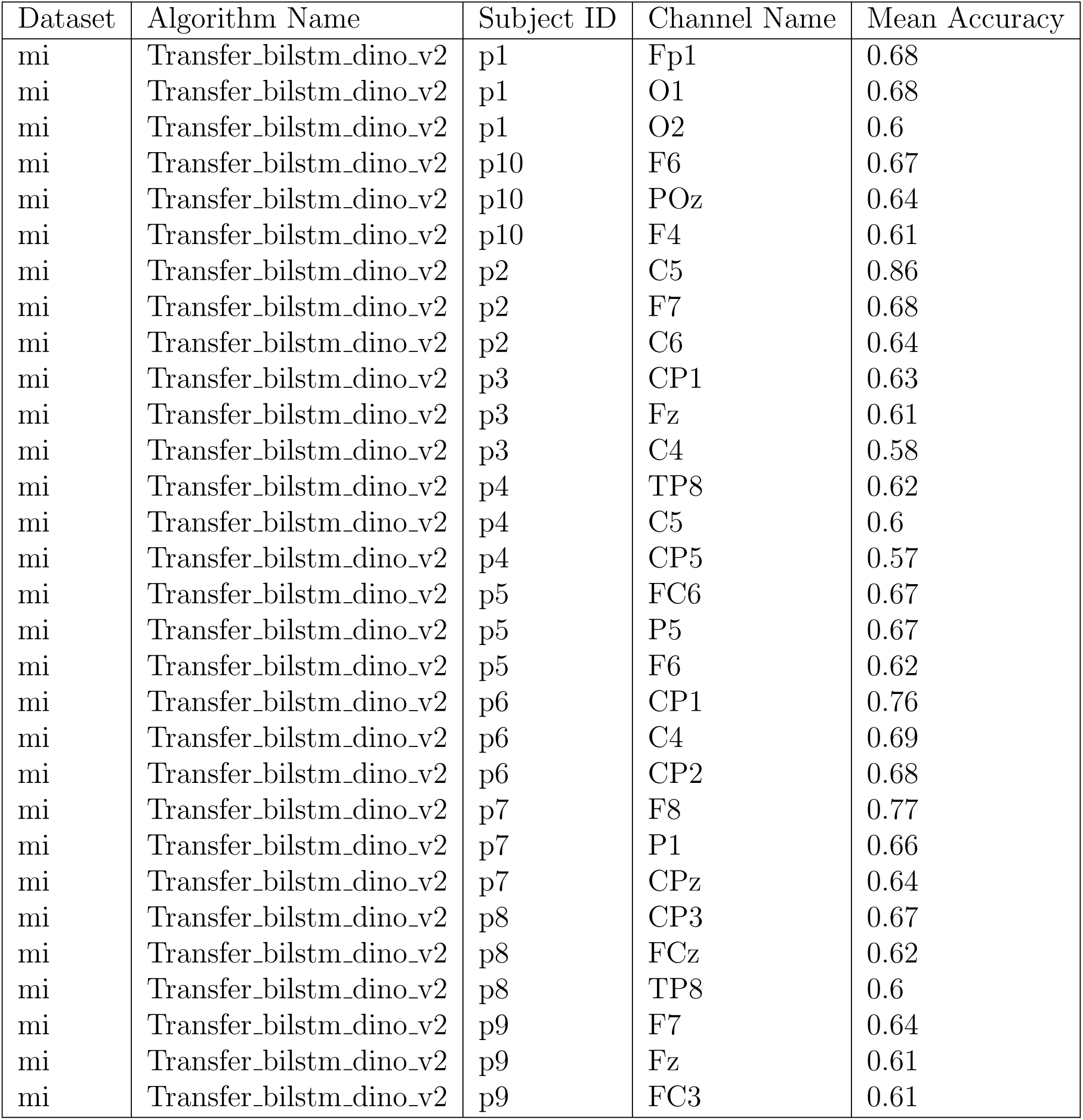
Summary of the classification accuracy for the top three channels for each subject, and dataset for EEGmmidb using Transfer_bilstm_dino_v2.

**Table S4:** Summary of the classification accuracy for the top three channels for each subject, and dataset for OpenMIIR using transfer_bilstm.

| Dataset | Algorithm Name | Subject ID | Channel Name | Mean Accuracy |
| --- | --- | --- | --- | --- |
| music | Transfer_bilstm | p1 | C3 | 0.61 |
| music | Transfer_bilstm | p1 | P1 | 0.59 |
| music | Transfer_bilstm | p1 | AF8 | 0.59 |
| music | Transfer_bilstm | p10 | F6 | 0.67 |
| music | Transfer_bilstm | p10 | POz | 0.64 |
| music | Transfer_bilstm | p10 | F4 | 0.61 |
| music | Transfer_bilstm | p2 | C5 | 0.86 |
| music | Transfer_bilstm | p2 | F7 | 0.68 |
| music | Transfer_bilstm | p2 | C6 | 0.64 |
| music | Transfer_bilstm | p3 | CP1 | 0.63 |
| music | Transfer_bilstm | p3 | Fz | 0.61 |
| music | Transfer_bilstm | p3 | C4 | 0.58 |
| music | Transfer_bilstm | p4 | F7 | 0.68 |
| music | Transfer_bilstm | p4 | Fpz | 0.65 |
| music | Transfer_bilstm | p4 | CP5 | 0.62 |
| music | Transfer_bilstm | p5 | F2 | 0.59 |
| music | Transfer_bilstm | p5 | FC6 | 0.58 |
| music | Transfer_bilstm | p5 | FC1 | 0.58 |
| music | Transfer_bilstm | p6 | F8 | 0.64 |
| music | Transfer_bilstm | p6 | AF8 | 0.61 |
| music | Transfer_bilstm | p6 | CP6 | 0.6 |
| music | Transfer_bilstm | p7 | CP5 | 0.58 |
| music | Transfer_bilstm | p7 | C4 | 0.58 |
| music | Transfer_bilstm | p7 | FC4 | 0.58 |
| music | Transfer_bilstm | p8 | CP3 | 0.67 |
| music | Transfer_bilstm | p8 | FCz | 0.62 |
| music | Transfer_bilstm | p8 | TP8 | 0.6 |
| music | Transfer_bilstm | p9 | PO4 | 0.64 |
| music | Transfer_bilstm | p9 | CP3 | 0.61 |
| music | Transfer_bilstm | p9 | FC1 | 0.6 |
| music | Transfer_bilstm | p11 | Fp1 | 0.66 |
| music | Transfer_bilstm | p11 | AF7 | 0.65 |
| music | Transfer_bilstm | p11 | AFz | 0.65 |
| music | Transfer_bilstm | p12 | P4 | 0.6 |
| music | Transfer_bilstm | p12 | F3 | 0.58 |
| music | Transfer_bilstm | p12 | FCz | 0.56 |

**Table S5:** Summary of the classification accuracy for the top three channels for each subject, and dataset for OpenMIIR using transfer_bilstm_dino_v2.

| Dataset | Algorithm Name | Subject ID | Channel Name | Mean Accuracy |
| --- | --- | --- | --- | --- |
| music | Transfer_bilstm_dino_v2 | p1 | FT7 | 0.62 |
| music | Transfer_bilstm_dino_v2 | p1 | AFz | 0.62 |
| music | Transfer_bilstm_dino_v2 | p1 | C3 | 0.61 |
| music | Transfer_bilstm_dino_v2 | p10 | F6 | 0.67 |
| music | Transfer_bilstm_dino_v2 | p10 | POz | 0.64 |
| music | Transfer_bilstm_dino_v2 | p10 | F4 | 0.61 |
| music | Transfer_bilstm_dino_v2 | p2 | C5 | 0.86 |
| music | Transfer_bilstm_dino_v2 | p2 | F7 | 0.68 |
| music | Transfer_bilstm_dino_v2 | p2 | C6 | 0.64 |
| music | Transfer_bilstm_dino_v2 | p3 | CP1 | 0.63 |
| music | Transfer_bilstm_dino_v2 | p3 | Fz | 0.61 |
| music | Transfer_bilstm_dino_v2 | p3 | C4 | 0.58 |
| music | Transfer_bilstm_dino_v2 | p4 | F6 | 0.64 |
| music | Transfer_bilstm_dino_v2 | p4 | F8 | 0.63 |
| music | Transfer_bilstm_dino_v2 | p4 | AFz | 0.62 |
| music | Transfer_bilstm_dino_v2 | p5 | TP7 | 0.65 |
| music | Transfer_bilstm_dino_v2 | p5 | F3 | 0.62 |
| music | Transfer_bilstm_dino_v2 | p5 | PO7 | 0.61 |
| music | Transfer_bilstm_dino_v2 | p6 | AF3 | 0.63 |
| music | Transfer_bilstm_dino_v2 | p6 | AFz | 0.6 |
| music | Transfer_bilstm_dino_v2 | p6 | C6 | 0.59 |
| music | Transfer_bilstm_dino_v2 | p7 | F5 | 0.57 |
| music | Transfer_bilstm_dino_v2 | p7 | O2 | 0.57 |
| music | Transfer_bilstm_dino_v2 | p7 | F6 | 0.57 |
| music | Transfer_bilstm_dino_v2 | p8 | CP3 | 0.67 |
| music | Transfer_bilstm_dino_v2 | p8 | FCz | 0.62 |
| music | Transfer_bilstm_dino_v2 | p8 | TP8 | 0.6 |
| music | Transfer_bilstm_dino_v2 | p9 | C1 | 0.66 |
| music | Transfer_bilstm_dino_v2 | p9 | FC1 | 0.65 |
| music | Transfer_bilstm_dino_v2 | p9 | CPz | 0.64 |
| music | Transfer_bilstm_dino_v2 | p11 | Fp1 | 0.71 |
| music | Transfer_bilstm_dino_v2 | p11 | AFz | 0.7 |
| music | Transfer_bilstm_dino_v2 | p11 | AF4 | 0.69 |
| music | Transfer_bilstm_dino_v2 | p12 | Pz | 0.62 |
| music | Transfer_bilstm_dino_v2 | p12 | P9 | 0.62 |
| music | Transfer_bilstm_dino_v2 | p12 | FT8 | 0.58 |

**Table S6:**
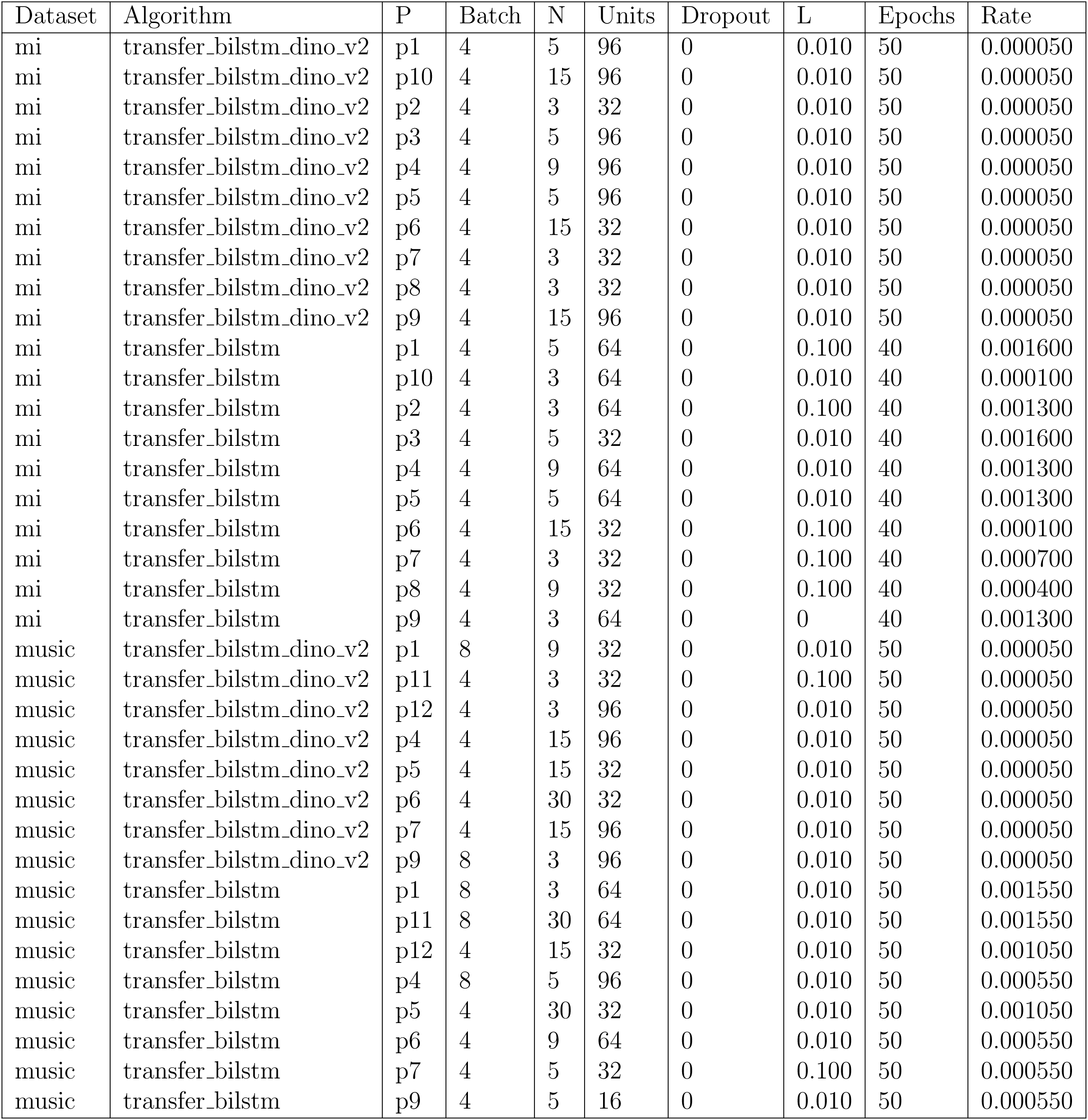
Results of our hyperparameter optimization process.

## Footnotes

1 This component will not be addressed in the scope of this paper.

2 https://mne.tools/stable/index.html

3 An example of one input matrix is displayed in Figure 2 in parts (c) and (d).

4 http://www.bci2000.org

## Notes

### Competing Interest Statement

The authors have declared no competing interest.

### Funding Statement

This study did not receive any funding

### Summary of Updates

order of the author list, I want to change Gailt to be the last.

## References

[1] Xiaoxi Wei. Transfer learning for non-invasive bci eeg brainwave decoding. 2024.

[2] Albina Li, Kanat Alimanov, Siamac Fazli, and Min-Ho Lee. Towards paradigm-independent brain computer interfaces. In 2020 8th International Winter Conference on Brain-Computer Interface (BCI), pages 1–6. IEEE, 2020.

[3] Donald Knuth. Knuth: Computers and typesetting.

[4] Muhammad F Mridha, Sujoy Chandra Das, Muhammad Mohsin Kabir, Aklima Akter Lima, Md Rashedul Islam, and Yutaka Watanobe. Brain-computer interface: Advancement and challenges. Sensors, 21(17):5746, 2021.

[5] Changkyun Im and Jong-Mo Seo. A review of electrodes for the electrical brain signal recording. Biomedical Engineering Letters, 6(3):104–112, 2016.

[6] Eleanor A Curran and Maria J Stokes. Learning to control brain activity: A review of the production and control of eeg components for driving brain–computer interface (bci) systems. Brain and cognition, 51(3):326–336, 2003.

[7] Maureen Clerc, Laurent Bougrain, and Fabien Lotte. Brain-computer interfaces 1: Methods and Perspectives. John Wiley & Sons, 2016.

[8] Amir Jalaly Bidgoly, Hamed Jalaly Bidgoly, and Zeynab Arezoumand. A survey on methods and challenges in eeg based authentication. Computers & Security, 93:101788, 2020.

[9] Lawrence M Ward. Synchronous neural oscillations and cognitive processes. Trends in cognitive sciences, 7(12):553–559, 2003.

[10] Yongtian He, David Eguren, José M Azorín, Robert G Grossman, Trieu Phat Luu, and Jose L Contreras-Vidal. Brain–machine interfaces for controlling lower-limb powered robotic systems. Journal of neural engineering, 15(2):021004, 2018.

[11] Wolfgang Ganglberger, Gerhard Gritsch, Manfred Martin Hartmann, Franz Fürbass, Hannes Perko, Ana M Skupch, and Tilmann Kluge. A comparison of rule-based and machine learning methods for classification of spikes in eeg. J. Commun., 12(10):589, 2017.

[12] Marzia De Lucia, Juan Fritschy, Peter Dayan, and David S Holder. A novel method for automated classification of epileptiform activity in the human electroencephalogram-based on independent component analysis. Medical & biological engineering & computing, 46(3):263–272, 2008.

[13] Lukas AW Gemein, Robin T Schirrmeister, Patryk szcz, Daniel Wilson, Joschka Boedecker, Andreas Schulze-Bonhage, Frank Hutter, and Tonio Ball. Machine-learning-based diagnostics of eeg pathology. NeuroImage, 220:117021, 2020.

[14] Hong Zeng, Chen Yang, Guojun Dai, Feiwei Qin, Jianhai Zhang, and Wanzeng Kong. Eeg classification of driver mental states by deep learning. Cognitive neurodynamics, 12(6):597–606, 2018.

[15] Zhichuan Tang, Chao Li, and Shouqian Sun. Single-trial eeg classification of motor imagery using deep convolutional neural networks. Optik, 130:11–18, 2017.

[16] Lawrence Ashley Farwell and Emanuel Donchin. Talking off the top of your head: toward a mental prosthesis utilizing event-related brain potentials. Electroencephalography and clinical Neurophysiology, 70(6):510–523, 1988.

[17] Faraz Akram, Seung Moo Han, and Tae-Seong Kim. An efficient word typing p300- bci system using a modified t9 interface and random forest classifier. Computers in biology and medicine, 56:30–36, 2015.

[18] Mohammad Norizadeh Cherloo, Amir Mohammad Mijani, Liang Zhan, and Mohammad Reza Daliri. A novel multiclass-based framework for p300 detection in bci matrix speller: Temporal eeg patterns of non-target trials vary based on their position to previous target stimuli. Engineering Applications of Artificial Intelligence, 123:106381, 2023.

[19] William O Tatum IV. Handbook of EEG interpretation. Springer Publishing Company, 2021.

[20] Nigel C Rogasch, Richard H Thomson, Faranak Farzan, Bernadette M Fitzgibbon, Neil W Bailey, Julio C Hernandez-Pavon, Zafiris J Daskalakis, and Paul B Fitzgerald. Removing artefacts from tms-eeg recordings using independent component analysis: importance for assessing prefrontal and motor cortex network properties. Neuroimage, 101:425–439, 2014.

[21] Atilla Kilicarslan, Robert G Grossman, and Jose Luis Contreras-Vidal. A robust adaptive denoising framework for real-time artifact removal in scalp eeg measurements. Journal of neural engineering, 13(2):026013, 2016.

[22] Yannick Roy, Hubert Banville, Isabela Albuquerque, Alexandre Gramfort, Tiago H Falk, and Jocelyn Faubert. Deep learning-based electroencephalography analysis: a systematic review. Journal of neural engineering, 16(5):051001, 2019.

[23] Alexander Craik, Yongtian He, and Jose L Contreras-Vidal. Deep learning for electroencephalogram (eeg) classification tasks: a review. Journal of neural engineering, 16(3):031001, 2019.

[24] Teng Ma, Hui Li, Hao Yang, Xulin Lv, Peiyang Li, Tiejun Liu, Dezhong Yao, and Peng Xu. The extraction of motion-onset vep bci features based on deep learning and compressed sensing. Journal of neuroscience methods, 275:80–92, 2017.

[25] Schubert R Carvalho, Iraquitan Cordeiro Filho, Damares Oliveira De Resende, Ana Carolina Siravenha, Cleidson RB De Souza, Henrique Debarba, Bruno D Gomes, and Ronan Boulic. A deep learning approach for classification of reaching targets from eeg images. In 2017 30th SIBGRAPI Conference on Graphics, Patterns and Images (SIBGRAPI), pages 178–184. IEEE, 2017.

[26] Guangyi Zhang, Vandad Davoodnia, Alireza Sepas-Moghaddam, Yaoxue Zhang, and Ali Etemad. Classification of hand movements from eeg using a deep attention-based lstm network. IEEE Sensors Journal, 20(6):3113–3122, 2019.

[27] Robin Tibor Schirrmeister, Jost Tobias Springenberg, Lukas Dominique Josef Fiederer, Martin Glasstetter, Katharina Eggensperger, Michael Tangermann, Frank Hutter, Wolfram Burgard, and Tonio Ball. Deep learning with convolutional neural networks for eeg decoding and visualization. Human brain mapping, 38(11):5391–5420, 2017.

[28] Md Zaved Iqubal Ahmed, Nidul Sinha, Souvik Phadikar, and Ebrahim Ghaderpour. Automated feature extraction on asmap for emotion classification using eeg. Sensors, 22(6):2346, 2022.

[29] Elnaz Lashgari, Dehua Liang, and Uri Maoz. Data augmentation for deep-learning-based electroencephalography. Journal of Neuroscience Methods, 346:108885, 2020.

[30] Fang Wang, Sheng-hua Zhong, Jianfeng Peng, Jianmin Jiang, and Yan Liu. Data augmentation for eeg-based emotion recognition with deep convolutional neural networks. In International conference on multimedia modeling, pages 82–93. Springer, 2018.

[31] Aigerim Keutayeva and Berdakh Abibullaev. Data constraints and performance optimization for transformer-based models in eeg-based brain-computer interfaces: A survey. IEEE Access, 2024.

[32] Qiqi Zhang and Ying Liu. Improving brain computer interface performance by data augmentation with conditional deep convolutional generative adversarial networks. arXiv preprint arXiv:1806.07108, 2018.

[33] Fabien Lotte, Laurent Bougrain, Andrzej Cichocki, Maureen Clerc, Marco Congedo, Alain Rakotomamonjy, and Florian Yger. A review of classification algorithms for eeg-based brain–computer interfaces: a 10 year update. Journal of neural engineering, 15(3):031005, 2018.

[34] Xiaoling Chen, Tianqing Li, Dongmei Liu, Shengcui Cheng, Dong Zhang, Juan Wang, Yuanyuan Zhang, and Ping Xie. Using transcranial magnetic stimulation to improve motor cortical excitability during motor imagery. IEEE Sensors Journal, 2024.

[35] Zhiwen Zhang, Feng Duan, Jordi Sole-Casals, Josep Dinares-Ferran, Andrzej Cichocki, Zhenglu Yang, and Zhe Sun. A novel deep learning approach with data augmentation to classify motor imagery signals. IEEE Access, 7:15945–15954, 2019.

[36] Gan Huang, Zhenxing Hu, Weize Chen, Shaorong Zhang, Zhen Liang, Linling Li, Li Zhang, and Zhiguo Zhang. M3cv: A multi-subject, multi-session, and multi-task database for eeg-based biometrics challenge. NeuroImage, 264:119666, 2022.

[37] Cheuk To Chung, Sharen Lee, Emma King, Tong Liu, Antonis A Armoundas, George Bazoukis, and Gary Tse. Clinical significance, challenges and limitations in using artificial intelligence for electrocardiography-based diagnosis. International journal of arrhythmia, 23(1):24, 2022.

[38] Jongmin Lee, Minju Kim, Dojin Heo, Jongsu Kim, Min-Ki Kim, Taejun Lee, Jong-woo Park, HyunYoung Kim, Minho Hwang, Laehyun Kim, et al. A comprehensive dataset for home appliance control using erp-based bcis with the application of inter-subject transfer learning. Frontiers in Human Neuroscience, 18:1320457, 2024.

[39] Goragod Pongthanisorn and Genci Capi. Genetic algorithm-based data optimization for efficient transfer learning in convolutional neural networks: A brain– machine interface implementation. Robotics, 13(1):14, 2024.

[40] Chia-Yen Yang, Pin-Chen Chen, and Wen-Chen Huang. Cross-domain transfer of eeg to eeg or ecg learning for cnn classification models. Sensors, 23(5):2458, 2023.

[41] Karl Weiss, Taghi M Khoshgoftaar, and DingDing Wang. A survey of transfer learning. Journal of Big data, 3(1):1–40, 2016.

[42] Ling Xiong, Nannan Li, Yi Luo, and Lei Chen. Sustainable development of electroencephalography materials and technology. SusMat, 4(2):e195, 2024.

[43] Jair Montoya-Martínez, Jonas Vanthornhout, Alexander Bertrand, and Tom Francart. Effect of number and placement of eeg electrodes on measurement of neural tracking of speech. Plos one, 16(2):e0246769, 2021.

[44] Vernon J Lawhern, Amelia J Solon, Nicholas R Waytowich, Stephen M Gordon, Chou P Hung, and Brent J Lance. Eegnet: a compact convolutional neural network for eeg-based brain–computer interfaces. Journal of neural engineering, 15(5):056013, 2018.

[45] Arunabha M Roy. An efficient multi-scale cnn model with intrinsic feature integration for motor imagery eeg subject classification in brain-machine interfaces. Biomedical Signal Processing and Control, 74:103496, 2022.

[46] Hongli Li, Man Ding, Ronghua Zhang, and Chunbo Xiu. Motor imagery eeg classification algorithm based on cnn-lstm feature fusion network. Biomedical signal processing and control, 72:103342, 2022.

[47] C Sidney Burrus. Wavelets and wavelet transforms. 2015.

[48] Nadia Mammone, Cosimo Ieracitano, and Francesco C Morabito. A deep cnn approach to decode motor preparation of upper limbs from time–frequency maps of eeg signals at source level. Neural Networks, 124:357–372, 2020.

[49] Mehmet Akif Ozdemir, Ozlem Karabiber Cura, and Aydin Akan. Epileptic eeg classification by using time-frequency images for deep learning. International journal of neural systems, 31(08):2150026, 2021.

[50] Xiuling Liu, Linyang Lv, Yonglong Shen, Peng Xiong, Jianli Yang, and Jing Liu. Multiscale space-time-frequency feature-guided multitask learning cnn for motor imagery eeg classification. Journal of Neural Engineering, 18(2):026003, 2021.

[51] Maxime Oquab, Timothée Darcet, Théo Moutakanni, Huy Vo, Marc Szafraniec, Vasil Khalidov, Pierre Fernandez, Daniel Haziza, Francisco Massa, Alaaeldin El-Nouby, et al. Dinov2: Learning robust visual features without supervision. arXiv preprint arXiv:2304.07193, 2023.

[52] Quentin Fournier, Gaétan Marceau Caron, and Daniel Aloise. A practical survey on faster and lighter transformers. ACM Computing Surveys, 55(14s):1–40, 2023.

[53] Shivarudhrappa Raghu, Natarajan Sriraam, Yasin Temel, Shyam Vasudeva Rao, and Pieter L Kubben. Eeg based multi-class seizure type classification using convolutional neural network and transfer learning. Neural Networks, 124:202–212, 2020.

[54] Jun Chin Ang, Andri Mirzal, Habibollah Haron, and Haza Nuzly Abdull Hamed. Supervised, unsupervised, and semi-supervised feature selection: a review on gene selection. IEEE/ACM transactions on computational biology and bioinformatics, 13(5):971–989, 2015.

[55] Abhilasha Nakra and Manoj Duhan. Deep neural network with harmony search based optimal feature selection of eeg signals for motor imagery classification. International Journal of Information Technology, 15(2):611–625, 2023.

[56] Jesús González, Julio Ortega, Miguel Damas, Pedro Martín-Smith, and John Q Gan. A new multi-objective wrapper method for feature selection–accuracy and stability analysis for bci. Neurocomputing, 333:407–418, 2019.

[57] Julio Ortega, Javier Asensio-Cubero, John Q Gan, and Andrés Ortiz. Classification of motor imagery tasks for bci with multiresolution analysis and multiobjective feature selection. BioMedical Engineering OnLine, 15:149–164, 2016.

[58] Tianqi Chen and Carlos Guestrin. Xgboost: A scalable tree boosting system. In Proceedings of the 22nd acm sigkdd international conference on knowledge discovery and data mining, pages 785–794, 2016.

[59] Sricheta Parui, Abhiroop Kumar Roshan Bajiya, Debasis Samanta, and Nishant Chakravorty. Emotion recognition from eeg signal using xgboost algorithm. In 2019 IEEE 16th India Council International Conference (INDICON), pages 1–4. IEEE, 2019.

[60] Fang Wang, Yu-Chu Tian, Xueying Zhang, and Fengyun Hu. An ensemble of xgboost models for detecting disorders of consciousness in brain injuries through eeg connectivity. Expert Systems with Applications, 198:116778, 2022.

[61] Hezam Albaqami, Ghulam Mubashar Hassan, Abdulhamit Subasi, and Amitava Datta. Automatic detection of abnormal eeg signals using wavelet feature extraction and gradient boosting decision tree. Biomedical Signal Processing and Control, 70:102957, 2021.

[62] Sepp Hochreiter and Jürgen Schmidhuber. Long short-term memory. Neural computation, 9(8):1735–1780, 1997.

[63] R Shashidhar, S Patilkulkarni, and SB Puneeth. Combining audio and visual speech recognition using lstm and deep convolutional neural network. International Journal of Information Technology, 14(7):3425–3436, 2022.

[64] Pedro Lopez-Rodriguez, Juan Gabriel Avina-Cervantes, Jose Luis Contreras-Hernandez, Rodrigo Correa, and Jose Ruiz-Pinales. Handwriting recognition based on 3d accelerometer data by deep learning. Applied Sciences, 12(13):6707, 2022.

[65] Sima Siami-Namini, Neda Tavakoli, and Akbar Siami Namin. The performance of lstm and bilstm in forecasting time series. In 2019 IEEE International Conference on Big Data (Big Data), pages 3285–3292. IEEE, 2019.

[66] Yousef Rezaei Tabar and Ugur Halici. A novel deep learning approach for classification of eeg motor imagery signals. Journal of neural engineering, 14(1):016003, 2016.

[67] Ping Wang, Aimin Jiang, Xiaofeng Liu, Jing Shang, and Li Zhang. Lstm-based eeg classification in motor imagery tasks. IEEE transactions on neural systems and rehabilitation engineering, 26(11):2086–2095, 2018.

[68] Jingcong Li, Zhu Liang Yu, Zhenghui Gu, Wei Wu, Yuanqing Li, and Lianwen Jin. A hybrid network for erp detection and analysis based on restricted boltzmann machine. IEEE Transactions on Neural Systems and Rehabilitation Engineering, 26(3):563–572, 2018.

[69] K Nandhini and G Tamilpavai. Hybrid cnn-lstm and modified wild horse herd model-based prediction of genome sequences for genetic disorders. Biomedical Signal Processing and Control, 78:103840, 2022.

[70] Jia Mian Tan, Haoran Liao, Wei Liu, Changjun Fan, Jincai Huang, Zhong Liu, and Junchi Yan. Hyperparameter optimization: Classics, acceleration, online, multi-objective, and tools. Mathematical Biosciences and Engineering, 21(6):6289–6335, 2024.

[71] Greeshma Sharma, Vishal Pandey, Ayush Chauhan, and Sushil Chandra. Deep learning approaches for electroencephalography (eeg)-based user response prediction. Artificial Intelligence Evolution, pages 226–233, 2023.

[72] Sebastian Stober, Avital Sternin, Adrian M Owen, and Jessica A Grahn. Towards music imagery information retrieval: Introducing the openmiir dataset of eeg recordings from music perception and imagination. In ISMIR, pages 763–769, 2015.

[73] Simanto Saha, Khondaker A Mamun, Khawza Ahmed, Raqibul Mostafa, Ganesh R Naik, Sam Darvishi, Ahsan H Khandoker, and Mathias Baumert. Progress in brain computer interface: Challenges and opportunities. Frontiers in systems neuroscience, 15:578875, 2021.

[74] Yi Gu and Lei Hua. A novel smart motor imagery intention human-computer interaction model using extreme learning machine and eeg signals. Frontiers in Neuroscience, 15:685119, 2021.

[75] Baoguo Xu, Wenlong Li, Xiaohang He, Zhiwei Wei, Dalin Zhang, Changcheng Wu, and Aiguo Song. Motor imagery based continuous teleoperation robot control with tactile feedback. Electronics, 9(1):174, 2020.

[76] Marti A. Hearst, Susan T Dumais, Edgar Osuna, John Platt, and Bernhard Scholkopf. Support vector machines. IEEE Intelligent Systems and their applications, 13(4):18–28, 1998.

[77] Mary Judith Antony, Baghavathi Priya Sankaralingam, Rakesh Kumar Mahendran, Akber Abid Gardezi, Muhammad Shafiq, Jin-Ghoo Choi, and Habib Hamam. Classification of eeg using adaptive svm classifier with csp and online recursive independent component analysis. Sensors, 22(19):7596, 2022.

[78] Maanvi Bhatnagar, Gauri Shanker Gupta, and Rakesh Kumar Sinha. Linear discriminant analysis classifies the eeg spectral features obtained from three class motor imagination. In 2018 2nd International Conference on Power, Energy and Environment: Towards Smart Technology (ICEPE), pages 1–6. IEEE, 2018.

[79] Yu-Te Wu, Tzu Hsuan Huang, Chun Yi Lin, Sheng Jia Tsai, and Po-Shan Wang. Classification of eeg motor imagery using support vector machine and convolutional neural network. In 2018 International Automatic Control Conference (CACS), pages 1–4. IEEE, 2018.

